# Statistical Analysis Plan for the Cardiac Magnetic Resonance GUIDEd Management of Mild-moderate Left Ventricular Systolic Dysfunction (CMR GUIDE) trial

**DOI:** 10.64898/2026.01.13.26344019

**Authors:** Laurent Billot, Jemima Gore, John Atherton, Colin Berry, Johannes Brachmann, Anand Ganesan, Graham Hillis, Werner Jung, Sanjay Prasad, Joseph Selvanayagam

## Abstract

The CMR Guide trial aims to determine whether the insertion of an Implantable Cardioverter Defibrillator based on the presence of ventricular scar/fibrosis will reduce sudden cardiac death or syncopal ventricular arrhythmia in patients with mild moderate left ventricle systolic dysfunction compared to standard care.

This statistical analysis plan pre-specifies the method of analysis for every outcome and key variables collected in the trial. The primary outcome is a composite of sudden cardiac death or haemodynamically significant ventricular arrhythmia defined as ventricular arrhythmia producing syncope (loss of consciousness) or associated with hypotension (SBP<90mmHg) except directly associated with device implant procedure.

The main analysis will consist of a Fine and Gray survival model of time to first occurrence of a primary outcome treating deaths other than sudden cardiac deaths as competing risks. The analysis plan also includes planned sensitivity analyses including covariate adjustments and subgroup analyses.

## 1 Administrative information

### 1.1 Study identifiers

- Protocol: Version 7 (25 June 2021)
- ClinicalTrials.gov register Identifier: <u>NCT01918215</u>

### 1.2 Revision history

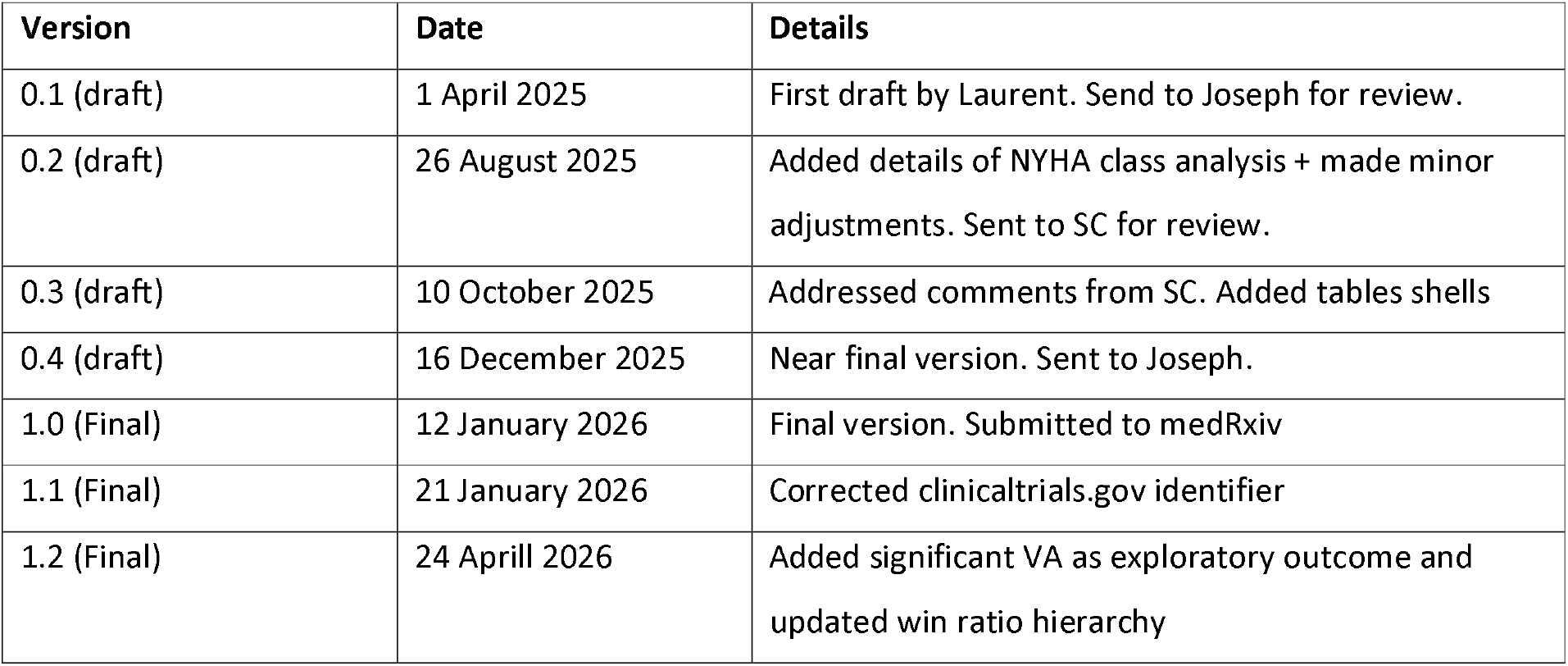

| Version | Date | Details |
| --- | --- | --- |
| 0.1 (draft) | 1 April 2025 | First draft by Laurent. Send to Joseph for review. |
| 0.2 (draft) | 26 August 2025 | Added details of NYHA class analysis + made minor adjustments. Sent to SC for review. |
| 0.3 (draft) | 10 October 2025 | Addressed comments from SC. Added tables shells |
| 0.4 (draft) | 16 December 2025 | Near final version. Sent to Joseph. |
| 1.0 (Final) | 12 January 2026 | Final version. Submitted to medRxiv |
| 1.1 (Final) | 21 January 2026 | Corrected clinicaltrials.gov identifier |
| 1.2 (Final) | 24 April 2026 | Added significant VA as exploratory outcome and updated win ratio hierarchy |

### 1.3 Contributors to the statistical analysis plan

#### 1.3.1 Roles and responsibilities

#### 1.3.2 Approvals

The undersigned have reviewed this plan and approve it as final. They find it to be consistent with the requirements of the protocol as it applies to their respective areas. They also find it to be compliant with ICH-E9 principles and, in particular, confirm that this analysis plan was developed in a completely blinded manner, i.e. without knowledge of the effect of the intervention(s) being assessed.

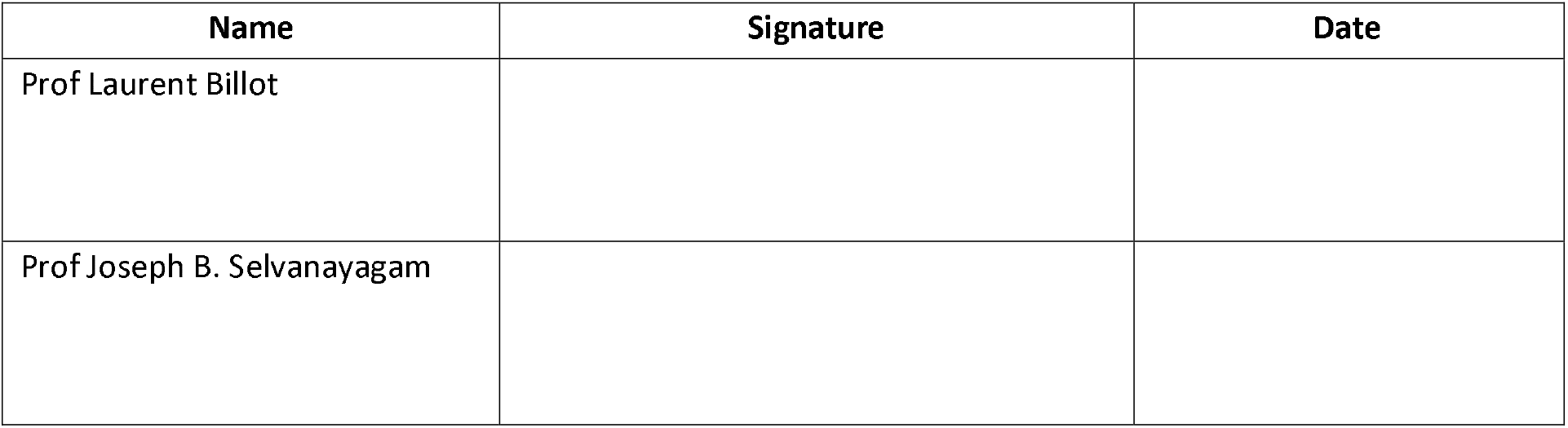

| Name | Signature | Date |
| --- | --- | --- |
| Prof Laurent Billot |  |  |
| Prof Joseph B. Selvanayagam |  |  |

## 2 Introduction

### 2.1 Study synopsis

The CMR Guide study is a prospective, multi-centre, randomised, controlled trial to determine whether the insertion of an Implantable Cardioverter Defibrillator (ICD) based on the presence of ventricular scar/fibrosis identified by Late Gadolinium Enhancement Cardiac Magnetic Resonance (LGE CMR) will reduce sudden cardiac death and/or syncopal ventricular arrhythmia in patients with mild moderate left ventricle (LV) systolic dysfunction compared to standard care.

The study will test the hypothesis that in a heterogeneous population of patients with mild-moderate left ventricular systolic dysfunction (with either ischemic or non-ischemic cardiomyopathy) and ventricular scar/fibrosis on cardiovascular magnetic resonance random assignment to ICD or implantable loop recorder (ILR) insertion results in different sudden cardiac death/syncope secondary to ventricular arrhythmia producing syncope (loss of consciousness) or associated with hypotension (SBP<90mmHg) rates.

The full study protocol is available in *Annals of Noninvasive Electrocardiology* [1].

### 2.2 Study population

#### 2.2.1 Inclusion Criteria

- Age ≥ 18 years old
- Coronary artery disease (CAD) or dilated cardiomyopathy (DCM) attributed to idiopathic, familial or chronic post myocarditis causes
- LVEF 36-50% by echocardiography, angiography, CMR or MUGA in the last 6 months (if a LGE CMR has been performed within the last 2 months then this can be used for inclusion and does not have to be repeated)
- On at least 3 months maximally tolerated doses of Angiotensin converting-enzyme (ACE) inhibitors (or angiotensin receptor blockers if intolerant of ACEi) and B blockers (If the dose has been changed considerably or a new drug started it will be at the PI’s discretion to deem eligibility)
- **Ischemic cardiomyopathy (ICM) patients**: CMR LVEF 36-50% and ventricular scar/fibrosis defined as LGE in 2 or more myocardial segments (17 segment AHA model) of transmural (>75%) hyper-enhancement
- **Nonischemic cardiomyopathy (NICM) patients:** CMR LVEF 36-50% and evidence of any scar/fibrosis on LGE
- Able and willing to comply with all pre-post- and follow-up testing and requirements

#### 2.2.2 Exclusion Criteria

- History of cardiac arrest or spontaneous or inducible sustained ventricular tachycardia (VT) or ventricular fibrillation (VF) unless occurring within 48 hours of an acute myocardial infarct (MI)
- Cardiomyopathy related to sarcoidosis
- Standard LGE CMR contraindications (e.g. severe claustrophobia, metal in-situ)
- Currently implanted permanent pacemaker/ICD and/or pacemaker/ICD lead
- Clinical indication for ICD or pacemaker or Cardiac Resynchronisation Therapy
- eGFR ≤30mls/min/1.73m2
- Recent MI (<40 days) or revascularization (<90 days)
- New York Heart Association HF functional class IV at the baseline visit
- Conditions associated with life expectancy <1 year
- Pregnancy or, in females of child-bearing potential, the non-use of accepted forms of contraception

### 2.3 Study interventions

#### 2.3.1 Randomisation

Participants are randomised using a computer-generated web-based randomisation schedule with stratification by site once the participant has their LGE CMR measurements verified by the Core Lab.

#### 2.3.2 Study treatment

##### Intervention (ICD) arm

Participants randomised to the ICD arm will have a single chamber ICD implanted and have the parameters programmed. Programming parameters will be standardised at the time of implant and represent a combination of the high rate and delayed therapy arms of the MADIT-RIT clinical trial both of which showed a reduction in the incidence of inappropriate shocks through delayed treatment in the VT zone and programming of high rate in the VF zone, while maintaining clinical effectiveness without increasing the rate of syncope. The monitor zone will allow correlation of rhythm in participants who present with syncope, allowing comparison with the ILR group. There will be two active tachycardia therapy zones and a further inactive monitor zone. Zones are determined by tachycardia cycle length (heart rate). There will be back up bradycardia pacing set at 40bpm VVI. A post implant chest X-ray is not mandated but will be at the discretion of the PI if clinically indicated.

##### Control (ILR) arm

Participants randomised to the ILR arm will have an implantable loop recorder implanted.

### 2.4 Outcomes

#### 2.4.1 Primary outcome

- Composite of sudden cardiac death (SCD) or haemodynamically significant ventricular arrhythmia (HSVA) defined as ventricular arrhythmia producing syncope (loss of consciousness) or associated with hypotension (SBP<90mmHg) except directly associated with device implant procedure.

#### 2.4.2 Secondary outcomes

- Sudden cardiac death
- Haemodynamically significant ventricular arrhythmia (HVA)
- All-cause mortality
- Cardiovascular mortality
- Quality of life assessed by Minnesota Living with Heart Failure Questionnaire (MLHFQ)
- Heart failure related hospitalisations

#### 2.4.3 Exploratory/other outcomes

- Sustained ventricular arrhythmia (VA) defined as a ventricular rhythm with a heart rate exceeding 100 beats per minute that either:

∘ persists for more than 30 seconds, OR
∘ results in appropriate therapy delivered by a programmed cardiac device, regardless of duration.

The arrhythmia must not be associated with syncope and must not be accompanied by documented systolic blood pressure <90 mmHg, as this constitutes the primary endpoint.

- Cardiovascular hospitalisations
- Change in New York Heart Association functional class
- EQ-5D-5L Questionnaire

### 2.5 Sample size

The trial is powered to detect a difference in the incidence of sudden cardiac death/syncope secondary to ventricular arrhythmia producing syncope (loss of consciousness) or associated with hypotension (SBP<90mmHg) based on a survival analysis of time to first event.

Assuming an annual event rate of 7.5% in the ILR group and a hazard ratio of 0.45, the trial requires at least 51 patients to experience a primary outcome event to reach 80% power (2-sided type-I error rate of 5%). We initially assumed uniform recruitment over 2 years and a total duration of about 5 years (average follow-up time of 4 years) which corresponds to the recruitment of around 250 patients. Assuming up to 20% of dropins (from ILR to ICD) and 10% of patients lost-to-follow up over the course of the trial, this led to a target sample size of around 428 patients.

A total of 353 patients were recruited over 7 years between mid-2015 and mid-2022. Follow-up is expected to be at least 3 years (until mid-2025) which should provide at least the same amount of power (more patient-years) as the initial study duration assumptions keeping the expected event rate and effect size the same. The figure below shows the expected power (y-axis) as a function of the yearly event rate in the control arm (x-axis) and expected hazard ratio (3 curves). With a hazard ratio of 0.45 (red curve) as initially assumed, the study should have at least 80% power for a yearly event rate in the control arm as low as 4%.

Power calculations displayed on the figure assume that 1% of patients in each arm are lost to follow-up every year and that 2% of patients allocated to the control arm cross over to the intervention (i.e. receive an ICD).

#### Power according to hazard ratio and event rate in the control arm

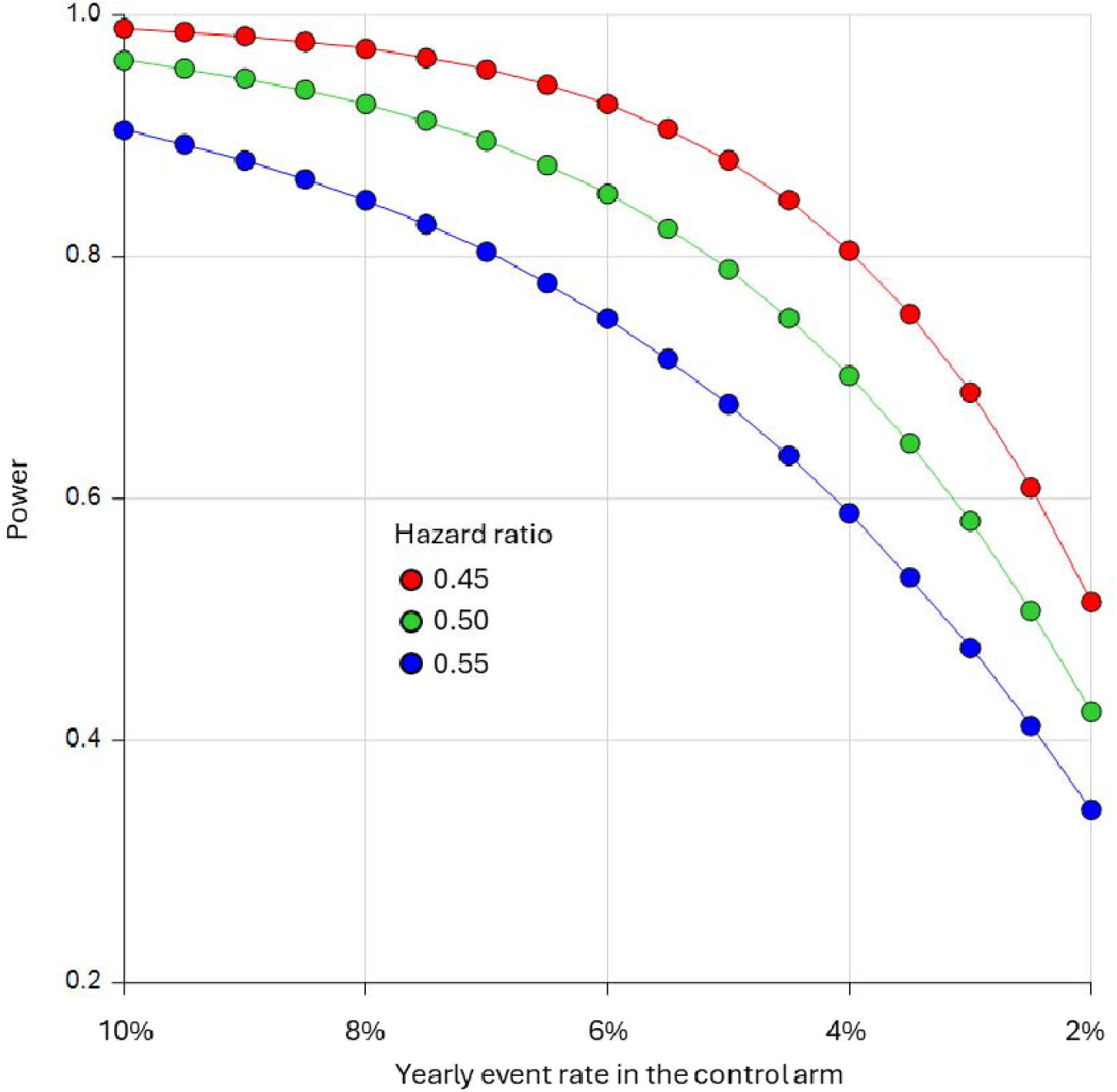

## 3 Statistical analysis

### 3.1 Software

Analyses will be conducted primarily using SAS Enterprise Guide (version 7.1 or above) and R (version 4.0.0 or above).

### 3.2 Interim analyses

A DSMB met to review safety data and overall progress approximately once a year. No formal interim efficacy analyses were undertaken; therefore, no adjustments are made to the final significance level.

### 3.3 Multiplicity adjustment

All tests are to be two-sided with a nominal level of α set at 5%. Analyses of the primary outcome (time to first occurrence of sudden cardiac death or haemodynamically significant ventricular arrhythmia) will be performed with an unadjusted type I error rate. Components of the primary outcome including (1) recurrent sudden cardiac death, and (2) Haemodynamically significant ventricular arrhythmia will also be analysed without applying any multiplicity adjustment. Instead, each will be analyzed regardless of statistical significance on the primary composite outcome but with only point estimates and 95% confidence intervals reported without significance tests.

For other secondary outcomes, we will control the family-wise error rate with a sequential Holm-Sidak correction [2]. This will be applied to the following secondary outcomes:

- All-cause mortality
- Quality of life assessed by Minnesota Living with Heart Failure Questionnaire (MLHFQ) overall score
- Heart failure related hospitalisations
- Cardiovascular mortality

Briefly, the approach consists of ordering all p-values from smallest to largest and then comparing them to an adjusted level of significance calculated as 1-(1-0.05)^1/C^, where C indicates the number of comparisons that remain. In the case of four secondary outcomes, the smallest p-value would be compared to 1-(1-0.05)^1/4^, the second p value to 1-(1-0.05)^1/3^, and so on, with the last one being compared to 1-(1-0.05) (i.e. 0.05). The sequential testing procedure stops as soon as a p-value fails to reach the corrected significance level. The multiplicity adjustment will only be applied to the primary analysis of each outcome. In case of sensitivity analyses (e.g. further adjustments), results will be presented as point estimates and 95% confidence intervals without significance tests. For MLHFQ, the adjustment will only apply to the analysis of the overall score.

No multiplicity adjustment will be applied to the analysis of other outcomes including compliance, vital signs and safety measures as well as exploratory outcomes (see Section 2.4.3). The corresponding results will be considered exploratory.

### 3.4 Data sets analysed

The primary hypothesis is whether a CMR-guided ICD implantation strategy leads to better outcomes than a standard approach. All analyses will therefore be performed on an intention-to-treat (ITT) basis by including all randomised patients according to their allocated group, regardless of treatment adherence and overall protocol compliance i.e. using a “Treatment Policy” strategy under the estimand terminology [3]. The ITT analysis set will be used to assess both efficacy and safety.

For events undergoing adjudication, only events that have been confirmed by the adjudication committee will be included in the analysis.

Exploratory analyses including “per-protocol” or “on-treatment” analyses may be undertaken later but will not be part of the main analysis and publication.

### 3.5 Handling of the ‘end of study’ visit

Given study participants enter the study at different times and are followed for various durations, the end of study (EOS) visit will occur at different times for different participants. For longitudinal and descriptive analyses by visit, EOS visits will be re-allocated according to the planned visit schedule. The allocation, based on the number of days between the date of randomization and the EOS visit, will be done as follows:

| Visit (month) | Window (days) | Visit (month) | Window (days) | Visit (month) | Window (days) |
| --- | --- | --- | --- | --- | --- |
| 6 | 92 to 274 | 30 | 824 to 1006 | 54 | 1556 to 1738 |
| 12 | 275 to 457 | 36 | 1007 to 1189 | 60 | 1739 to 1921 |
| 18 | 458 to 640 | 42 | 1190 to 1372 | 66 | 1922 to 2104 |
| 24 | 641 to 823 | 48 | 1373 to 1555 | 72 | 2105 to 2287 |

In case of a reallocated EOS visit falling within a window already containing a scheduled visit, we will retain the value from the previous scheduled visit, unless it is missing.

### 3.6 Subject disposition

The flow of patients through the trial will be displayed in a CONSORT [4] (Consolidated Standards of Reporting Trials) diagram. The report will include the following: the number of CMR screened patients who met study inclusion criteria and the number of patients who were included; and reasons for exclusion of non-included patients. A table will summarize the data completeness and study withdrawal.

### 3.7 Patient characteristics and baseline comparisons

Description of the baseline characteristics will be presented by treatment group as outlined in Appendix 1 (Table 1). Discrete variables will be summarised by frequencies and percentages. Percentages will be calculated according to the number of patients for whom data are available. Continuous variables will be summarised using mean and SD, and median and interquartile range (Q1-Q3). Baseline measures for all patients will be tabulated for the variables listed below:

**Table 1.** **Subject disposition**

|  | ICD | ILR | Total |
| --- | --- | --- | --- |
| Screened | xxx | xxx | xxx |
| Randomised | xxx (100%) | xxx (100%) | xxx (100%) |
| Device inserted | xxx (xx.x%) | xxx (xx.x%) | xxx (xx.x%) |
| Completed study | xxx (xx.x%) | xxx (xx.x%) | xxx (xx.x%) |
| Withdrawn* | xxx (xx.x%) | xxx (xx.x%) | xxx (xx.x%) |
| Death | xxx (xx.x%) | xxx (xx.x%) | xxx (xx.x%) |
| Patient or family request | xxx (xx.x%) | xxx (xx.x%) | xxx (xx.x%) |
| Patient Lost to follow up | xxx (xx.x%) | xxx (xx.x%) | xxx (xx.x%) |
| Patient non-compliance | xxx (xx.x%) | xxx (xx.x%) | xxx (xx.x%) |
| Patient participation termination by Investigator | xxx (xx.x%) | xxx (xx.x%) | xxx (xx.x%) |
| AE | xxx (xx.x%) | xxx (xx.x%) | xxx (xx.x%) |
| SAE | xxx (xx.x%) | xxx (xx.x%) | xxx (xx.x%) |
| CMR showed LVEF>50% | xxx (xx.x%) | xxx (xx.x%) | xxx (xx.x%) |
| Other | xxx (xx.x%) | xxx (xx.x%) | xxx (xx.x%) |
| Unknown | xxx (xx.x%) | xxx (xx.x%) | xxx (xx.x%) |
|  | xxx (xx.x%) | xxx (xx.x%) | xxx (xx.x%) |
| Month 3 visit performed | xxx (xx.x%) | xxx (xx.x%) | xxx (xx.x%) |
| Month 6 visit performed | xxx (xx.x%) | xxx (xx.x%) | xxx (xx.x%) |
| Month 12 visit performed | xxx (xx.x%) | xxx (xx.x%) | xxx (xx.x%) |
| Month 24 visit performed | xxx (xx.x%) | xxx (xx.x%) | xxx (xx.x%) |
| Month 36 visit performed | xxx (xx.x%) | xxx (xx.x%) | xxx (xx.x%) |
| Month 48 visit performed | xxx (xx.x%) | xxx (xx.x%) | xxx (xx.x%) |
| Month 60 visit performed | xxx (xx.x%) | xxx (xx.x%) | xxx (xx.x%) |
| Month 72 visit performed | xxx (xx.x%) | xxx (xx.x%) | xxx (xx.x%) |
| Month 84 visit performed | xxx (xx.x%) | xxx (xx.x%) | xxx (xx.x%) |
| Month 96 visit performed | xxx (xx.x%) | xxx (xx.x%) | xxx (xx.x%) |
| Month 108 visit performed | xxx (xx.x%) | xxx (xx.x%) | xxx (xx.x%) |
| End of study | xxx (xx.x%) | xxx (xx.x%) | xxx (xx.x%) |

**Notes:**
*Except for withdrawal reasons, the denominator is the number of randomised patients. For the distribution of withdrawal reasons, the denominator is the number of patients withdrawn.*
*\*All deaths may not be accompanied by a study withdrawal, hence the number of deaths as a withdrawal reason may be smaller than recorded deaths.*

**Table 2.**
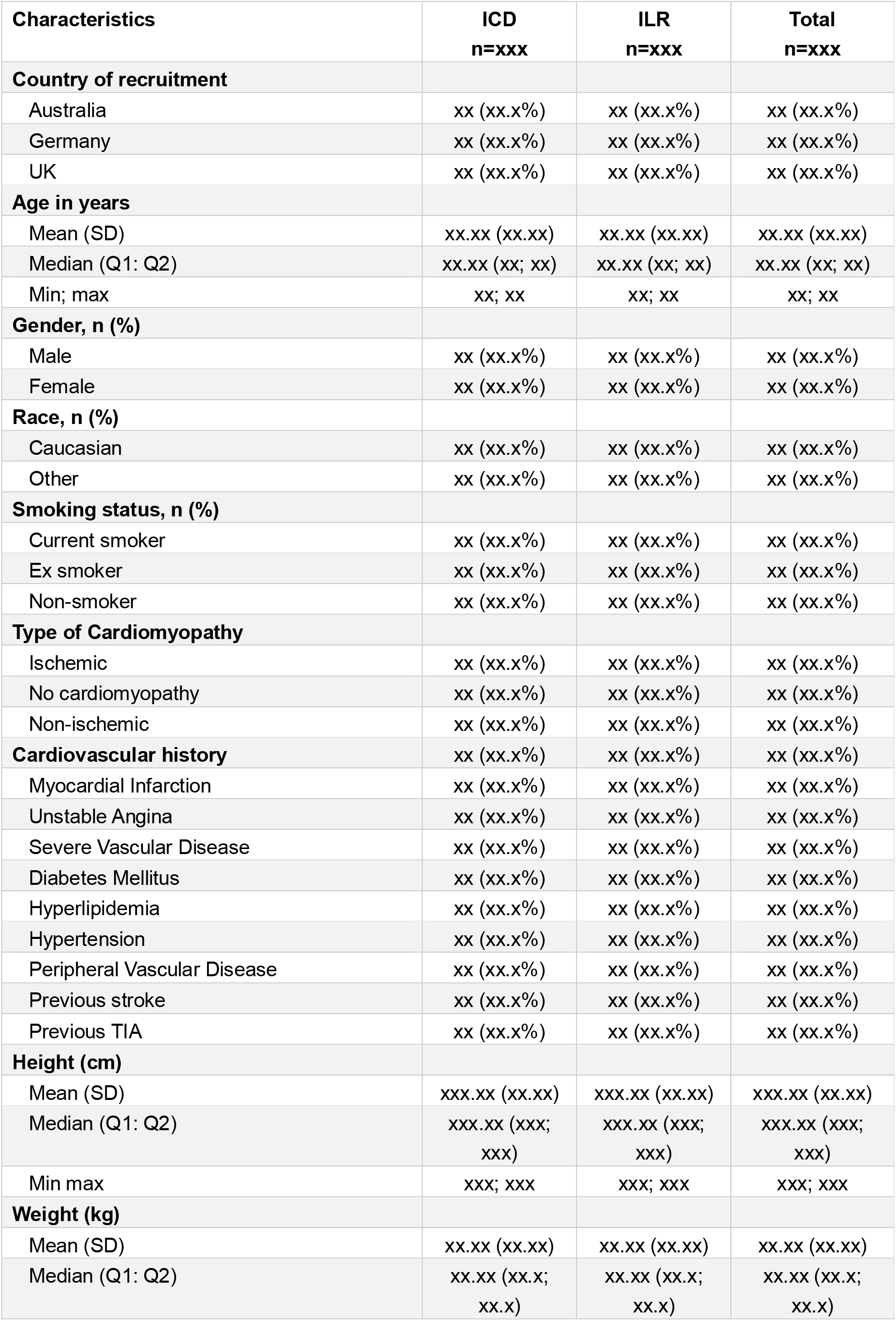
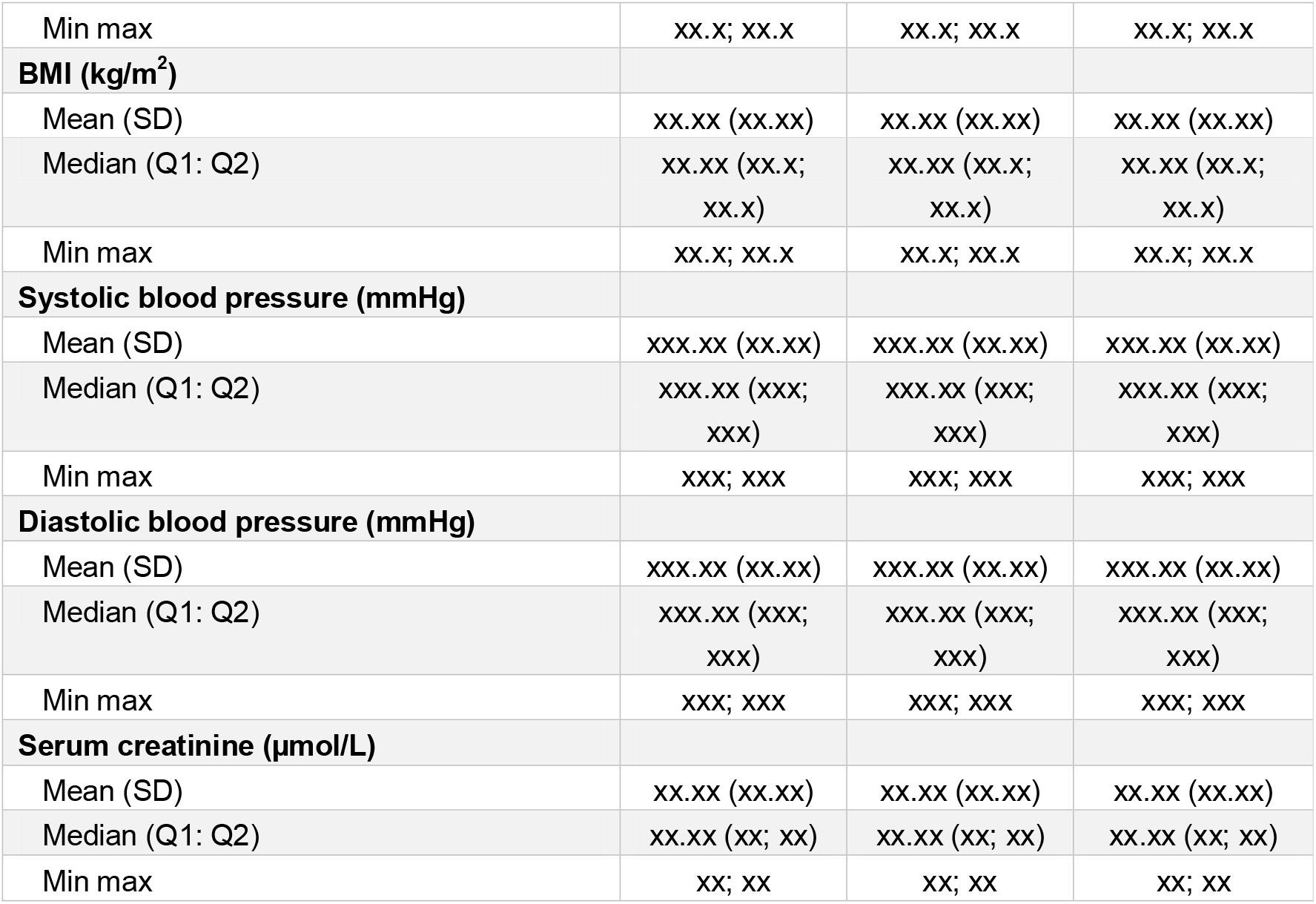
Key baseline characteristics.

| **Characteristics** | **ICD**<br>**n=xxx** | **ILR**<br>**n=xxx** | **Total**<br>**n=xxx** |
| --- | --- | --- | --- |
| **Country of recruitment** |  |  |  |
| Australia | xx (xx.x%) | xx (xx.x%) | xx (xx.x%) |
| Germany | xx (xx.x%) | xx (xx.x%) | xx (xx.x%) |
| UK | xx (xx.x%) | xx (xx.x%) | xx (xx.x%) |
| **Age in years** |  |  |  |
| Mean (SD) | xx.xx (xx.xx) | xx.xx (xx.xx) | xx.xx (xx.xx) |
| Median (Q1: Q2) | xx.xx (xx; xx) | xx.xx (xx; xx) | xx.xx (xx; xx) |
| Min; max | xx; xx | xx; xx | xx; xx |
| **Gender, n (%)** |  |  |  |
| Male | xx (xx.x%) | xx (xx.x%) | xx (xx.x%) |
| Female | xx (xx.x%) | xx (xx.x%) | xx (xx.x%) |
| **Race, n (%)** |  |  |  |
| Caucasian | xx (xx.x%) | xx (xx.x%) | xx (xx.x%) |
| Other | xx (xx.x%) | xx (xx.x%) | xx (xx.x%) |
| **Smoking status, n (%)** |  |  |  |
| Current smoker | xx (xx.x%) | xx (xx.x%) | xx (xx.x%) |
| Ex smoker | xx (xx.x%) | xx (xx.x%) | xx (xx.x%) |
| Non-smoker | xx (xx.x%) | xx (xx.x%) | xx (xx.x%) |
| **Type of Cardiomyopathy** |  |  |  |
| Ischemic | xx (xx.x%) | xx (xx.x%) | xx (xx.x%) |
| No cardiomyopathy | xx (xx.x%) | xx (xx.x%) | xx (xx.x%) |
| Non-ischemic | xx (xx.x%) | xx (xx.x%) | xx (xx.x%) |
| **Cardiovascular history** | xx (xx.x%) | xx (xx.x%) | xx (xx.x%) |
| Myocardial Infarction | xx (xx.x%) | xx (xx.x%) | xx (xx.x%) |
| Unstable Angina | xx (xx.x%) | xx (xx.x%) | xx (xx.x%) |
| Severe Vascular Disease | xx (xx.x%) | xx (xx.x%) | xx (xx.x%) |
| Diabetes Mellitus | xx (xx.x%) | xx (xx.x%) | xx (xx.x%) |
| Hyperlipidemia | xx (xx.x%) | xx (xx.x%) | xx (xx.x%) |
| Hypertension | xx (xx.x%) | xx (xx.x%) | xx (xx.x%) |
| Peripheral Vascular Disease | xx (xx.x%) | xx (xx.x%) | xx (xx.x%) |
| Previous stroke | xx (xx.x%) | xx (xx.x%) | xx (xx.x%) |
| Previous TIA | xx (xx.x%) | xx (xx.x%) | xx (xx.x%) |
| **Height (cm)** |  |  |  |
| Mean (SD) | xxx.xx (xx.xx) | xxx.xx (xx.xx) | xxx.xx (xx.xx) |
| Median (Q1: Q2) | xxx.xx (xxx;<br>xxx) | xxx.xx (xxx;<br>xxx) | xxx.xx (xxx;<br>xxx) |
| Min max | xxx; xxx | xxx; xxx | xxx; xxx |
| **Weight (kg)** |  |  |  |
| Mean (SD) | xx.xx (xx.xx) | xx.xx (xx.xx) | xx.xx (xx.xx) |
| Median (Q1: Q2) | xx.xx (xx.x;<br>xx.x) | xx.xx (xx.x;<br>xx.x) | xx.xx (xx.x;<br>xx.x) |
| Min max | xx.x; xx.x | xx.x; xx.x | xx.x; xx.x |
| **BMI (kg/m<sup>2</sup>)** |  |  |  |
| Mean (SD) | xx.xx (xx.xx) | xx.xx (xx.xx) | xx.xx (xx.xx) |
| Median (Q1: Q2) | xx.xx (xx.x;<br>xx.x) | xx.xx (xx.x;<br>xx.x) | xx.xx (xx.x;<br>xx.x) |
| Min max | xx.x; xx.x | xx.x; xx.x | xx.x; xx.x |
| **Systolic blood pressure (mmHg)** |  |  |  |
| Mean (SD) | xxx.xx (xx.xx) | xxx.xx (xx.xx) | xxx.xx (xx.xx) |
| Median (Q1: Q2) | xxx.xx (xxx;<br>xxx) | xxx.xx (xxx;<br>xxx) | xxx.xx (xxx;<br>xxx) |
| Min max | xxx; xxx | xxx; xxx | xxx; xxx |
| **Diastolic blood pressure (mmHg)** |  |  |  |
| Mean (SD) | xxx.xx (xx.xx) | xxx.xx (xx.xx) | xxx.xx (xx.xx) |
| Median (Q1: Q2) | xxx.xx (xxx;<br>xxx) | xxx.xx (xxx;<br>xxx) | xxx.xx (xxx;<br>xxx) |
| Min max | xxx; xxx | xxx; xxx | xxx; xxx |
| **Serum creatinine (μmol/L)** |  |  |  |
| Mean (SD) | xx.xx (xx.xx) | xx.xx (xx.xx) | xx.xx (xx.xx) |
| Median (Q1: Q2) | xx.xx (xx; xx) | xx.xx (xx; xx) | xx.xx (xx; xx) |
| Min max | xx; xx | xx; xx | xx; xx |

**Table 3:** Baseline medications.

|  | ICD | ILR | Total |
| --- | --- | --- | --- |
|  | N=XXX | N=XXX | N=XXX |
| Additional Cardiac Medications | xx/xxx (%) | xx/xxx (%) | xx/xxx (%) |
| Aldosterone Receptor Antagonists | xx/xxx (%) | xx/xxx (%) | xx/xxx (%) |
| Angiotensin Converting Enzyme inhibitors | xx/xxx (%) | xx/xxx (%) | xx/xxx (%) |
| Angiotensin II Receptor Blockers | xx/xxx (%) | xx/xxx (%) | xx/xxx (%) |
| Angiotensin receptor neprilysin inhibitors | xx/xxx (%) | xx/xxx (%) | xx/xxx (%) |
| Anti-reflux medications | xx/xxx (%) | xx/xxx (%) | xx/xxx (%) |
| Antiarrhythmic therapy | xx/xxx (%) | xx/xxx (%) | xx/xxx (%) |
| Antibiotics and antimicrobials | xx/xxx (%) | xx/xxx (%) | xx/xxx (%) |
| Anticoagulation therapy | xx/xxx (%) | xx/xxx (%) | xx/xxx (%) |
| Antilipid | xx/xxx (%) | xx/xxx (%) | xx/xxx (%) |
| Antiplatelet therapy | xx/xxx (%) | xx/xxx (%) | xx/xxx (%) |
| Beta Blockers | xx/xxx (%) | xx/xxx (%) | xx/xxx (%) |
| Ca Channel blockers | xx/xxx (%) | xx/xxx (%) | xx/xxx (%) |
| Digoxin | xx/xxx (%) | xx/xxx (%) | xx/xxx (%) |
| Diuretics | xx/xxx (%) | xx/xxx (%) | xx/xxx (%) |
| Hypoglycaemic agents | xx/xxx (%) | xx/xxx (%) | xx/xxx (%) |
| Inotropes | xx/xxx (%) | xx/xxx (%) | xx/xxx (%) |
| Pain relievers | xx/xxx (%) | xx/xxx (%) | xx/xxx (%) |
| SGLT2 inhibitors | xx/xxx (%) | xx/xxx (%) | xx/xxx (%) |
| Vitamin and Mineral supplements | xx/xxx (%) | xx/xxx (%) | xx/xxx (%) |
| Unknown | xx/xxx (%) | xx/xxx (%) | xx/xxx (%) |
| Other | xx/xxx (%) | xx/xxx (%) | xx/xxx (%) |

**Table 4:** Concomitant medications during follow-up.

|  | **ICD**<br>**N=XXX** | **ILR**<br>**N=XXX** | **Total**<br>**N=XXX** |
| --- | --- | --- | --- |
| Additional Cardiac Medications | xx/xxx (%) | xx/xxx (%) | xx/xxx (%) |
| Aldosterone Receptor Antagonists | xx/xxx (%) | xx/xxx (%) | xx/xxx (%) |
| Angiotensin Converting Enzyme inhibitors | xx/xxx (%) | xx/xxx (%) | xx/xxx (%) |
| Angiotensin II Receptor Blockers | xx/xxx (%) | xx/xxx (%) | xx/xxx (%) |
| Angiotensin receptor neprilysin inhibitors | xx/xxx (%) | xx/xxx (%) | xx/xxx (%) |
| Anti-reflux medications | xx/xxx (%) | xx/xxx (%) | xx/xxx (%) |
| Antiarrhythmic therapy | xx/xxx (%) | xx/xxx (%) | xx/xxx (%) |
| Antibiotics and antimicrobials | xx/xxx (%) | xx/xxx (%) | xx/xxx (%) |
| Anticoagulation therapy | xx/xxx (%) | xx/xxx (%) | xx/xxx (%) |
| Antilipid | xx/xxx (%) | xx/xxx (%) | xx/xxx (%) |
| Antiplatelet therapy | xx/xxx (%) | xx/xxx (%) | xx/xxx (%) |
| Beta Blockers | xx/xxx (%) | xx/xxx (%) | xx/xxx (%) |
| Ca Channel blockers | xx/xxx (%) | xx/xxx (%) | xx/xxx (%) |
| Digoxin | xx/xxx (%) | xx/xxx (%) | xx/xxx (%) |
| Diuretics | xx/xxx (%) | xx/xxx (%) | xx/xxx (%) |
| Hypoglycaemic agents | xx/xxx (%) | xx/xxx (%) | xx/xxx (%) |
| Inotropes | xx/xxx (%) | xx/xxx (%) | xx/xxx (%) |
| Pain relievers | xx/xxx (%) | xx/xxx (%) | xx/xxx (%) |
| SGLT2 inhibitors | xx/xxx (%) | xx/xxx (%) | xx/xxx (%) |
| Vitamin and Mineral supplements | xx/xxx (%) | xx/xxx (%) | xx/xxx (%) |
| Unknown | xx/xxx (%) | xx/xxx (%) | xx/xxx (%) |
| Other | xx/xxx (%) | xx/xxx (%) | xx/xxx (%) |

**Table 5.** Compliance and protocol deviations.

| Type of protocol deviation | ICD<br>n=xxx | ILR<br>n=xxx | Total<br>n=xxx |
| --- | --- | --- | --- |
| Inclusion/Exclusion Criteria not met | xx (xx.x%) | xx (xx.x%) | xx (xx.x%) |
| Visit outside window | xx (xx.x%) | xx (xx.x%) | xx (xx.x%) |
| Visit not done | xx (xx.x%) | xx (xx.x%) | xx (xx.x%) |
| Required tests not done (e.g. bloods, MRI) | xx (xx.x%) | xx (xx.x%) | xx (xx.x%) |
| Data corrupted | xx (xx.x%) | xx (xx.x%) | xx (xx.x%) |
| Received the wrong randomised device* | xx (xx.x%) | xx (xx.x%) | xx (xx.x%) |
| Did not receive or deactivated the randomised device | xx (xx.x%) | xx (xx.x%) | xx (xx.x%) |
| Other | xx (xx.x%) | xx (xx.x%) | xx (xx.x%) |

\* Patients randomised to ILR who receive an ICD and patients randomised to ICD who received ILR

**Table 5.**
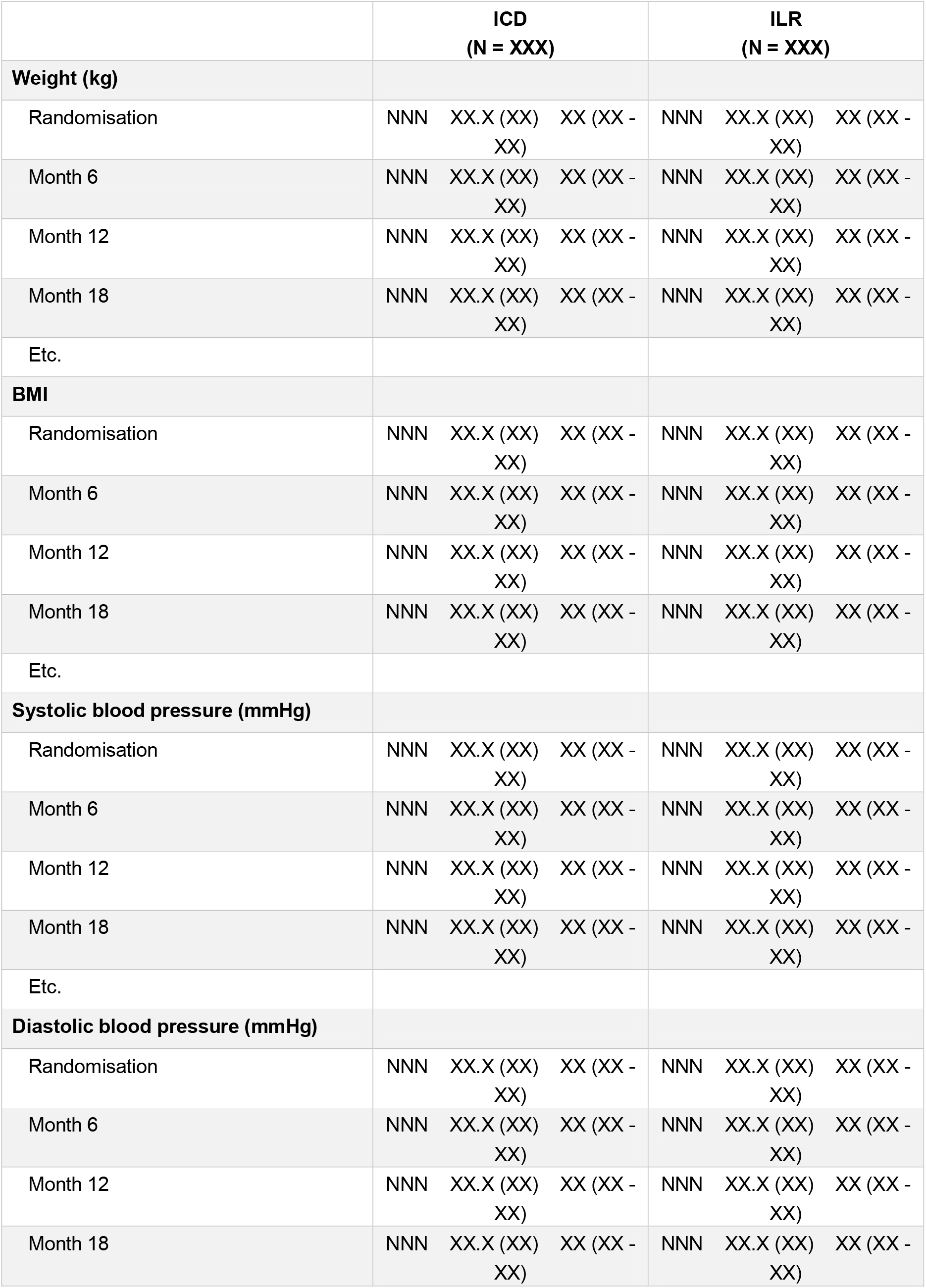
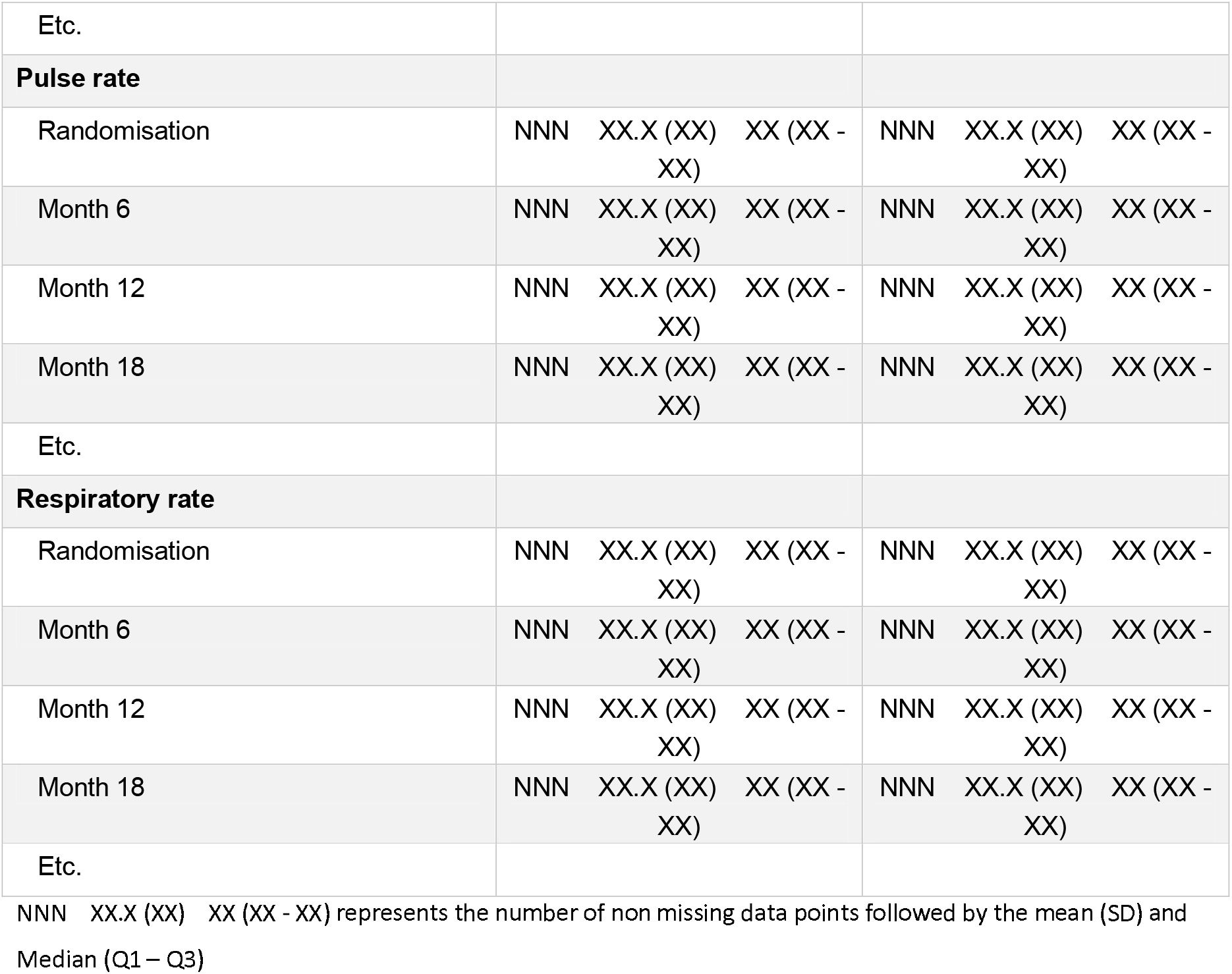
Vital signs by visit.

**Table 6.**
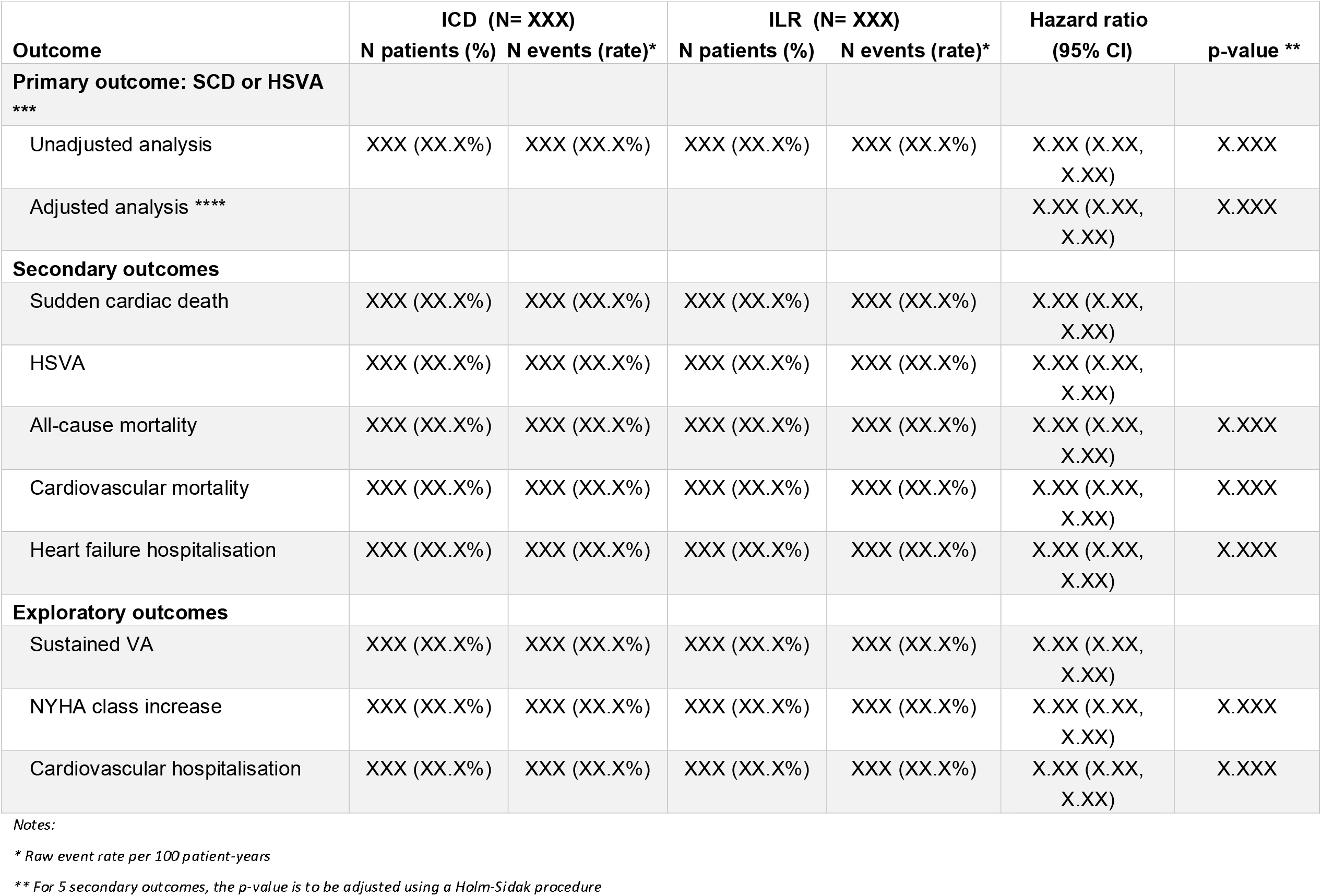
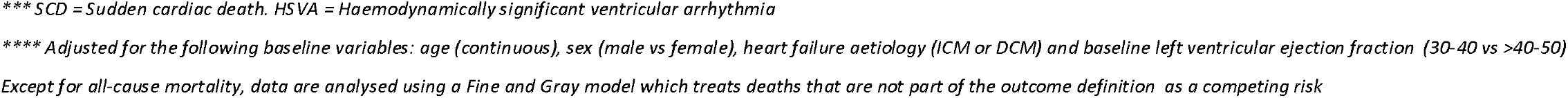
Survival analysis of primary and secondary outcomes.

| Outcome | ICD (N= XXX) |  | ILR (N= XXX) |  | Hazard ratio<br>(95% CI) | p-value ** |
| --- | --- | --- | --- | --- | --- | --- |
|  | N patients (%) | N events (rate)* | N patients (%) | N events (rate)* |  |  |
| **Primary outcome: SCD or HSVA**<br>**\*\*\*** |  |  |  |  |  |  |
| Unadjusted analysis | XXX (XX.X%) | XXX (XX.X%) | XXX (XX.X%) | XXX (XX.X%) | X.XX (X.XX,<br>X.XX) | X.XXX |
| Adjusted analysis **** |  |  |  |  | X.XX (X.XX,<br>X.XX) | X.XXX |
| **Secondary outcomes** |  |  |  |  |  |  |
| Sudden cardiac death | XXX (XX.X%) | XXX (XX.X%) | XXX (XX.X%) | XXX (XX.X%) | X.XX (X.XX,<br>X.XX) |  |
| HSVA | XXX (XX.X%) | XXX (XX.X%) | XXX (XX.X%) | XXX (XX.X%) | X.XX (X.XX,<br>X.XX) |  |
| All-cause mortality | XXX (XX.X%) | XXX (XX.X%) | XXX (XX.X%) | XXX (XX.X%) | X.XX (X.XX,<br>X.XX) | X.XXX |
| Cardiovascular mortality | XXX (XX.X%) | XXX (XX.X%) | XXX (XX.X%) | XXX (XX.X%) | X.XX (X.XX,<br>X.XX) | X.XXX |
| Heart failure hospitalisation | XXX (XX.X%) | XXX (XX.X%) | XXX (XX.X%) | XXX (XX.X%) | X.XX (X.XX,<br>X.XX) | X.XXX |
| **Exploratory outcomes** |  |  |  |  |  |  |
| Sustained VA | XXX (XX.X%) | XXX (XX.X%) | XXX (XX.X%) | XXX (XX.X%) | X.XX (X.XX,<br>X.XX) |  |
| NYHA class increase | XXX (XX.X%) | XXX (XX.X%) | XXX (XX.X%) | XXX (XX.X%) | X.XX (X.XX,<br>X.XX) | X.XXX |
| Cardiovascular hospitalisation | XXX (XX.X%) | XXX (XX.X%) | XXX (XX.X%) | XXX (XX.X%) | X.XX (X.XX,<br>X.XX) | X.XXX |

Notes:
\* Raw event rate per 100 patient-years
\*\* For 5 secondary outcomes, the p-value is to be adjusted using a Holm-Sidak procedure
\*\*\* SCD = Sudden cardiac death. HSVA = Haemodynamically significant ventricular arrhythmia
\*\*\*\* Adjusted for the following baseline variables: age (continuous), sex (male vs female), heart failure aetiology (ICM or DCM) and baseline left ventricular ejection fraction (30-40 vs >40-50)
Except for all-cause mortality, data are analysed using a Fine and Gray model which treats deaths that are not part of the outcome definition as a competing risk

**Table 7.** Sensitivity analyses of survival outcomes using cause-specific Cox models.

| **Outcome** | **Hazard ratio**<br>**(95% CI)** |
| --- | --- |
| Primary outcome: SCD or HSVA * | X.XX (X.XX, X.XX) |
| Sudden cardiac death | X.XX (X.XX, X.XX) |
| HSVA | X.XX (X.XX, X.XX) |
| Cardiovascular mortality | X.XX (X.XX, X.XX) |
| Heart failure hospitalisation | X.XX (X.XX, X.XX) |
| Sustained VA | X.XX (X.XX, X.XX) |
| NYHA class increase | X.XX (X.XX, X.XX) |
| Cardiovascular hospitalisation | X.XX (X.XX, X.XX) |

\* SCD = Sudden cardiac death. HSVA = Haemodynamically significant ventricular arrhythmia

**Table 8:**
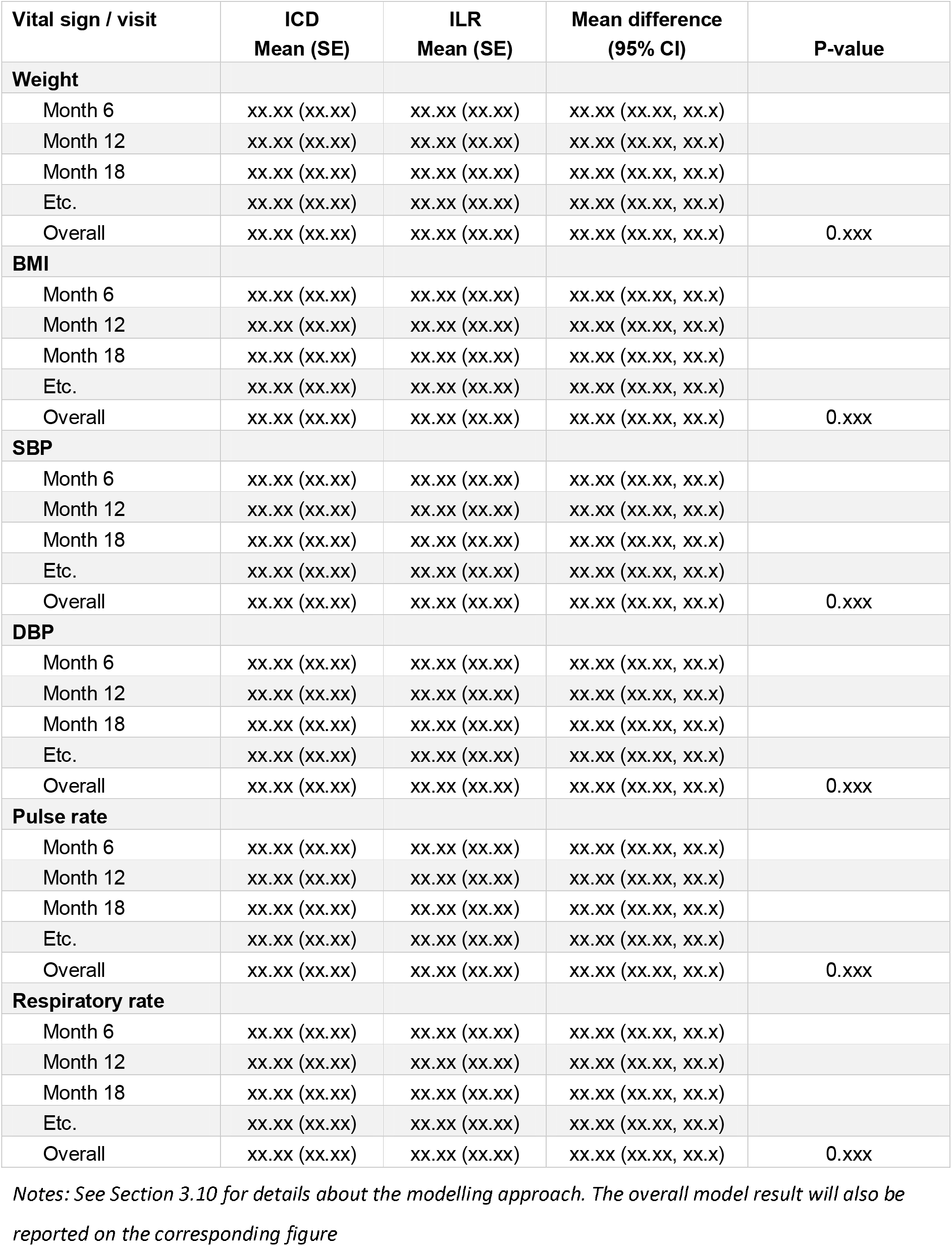
Analysis of vital signs.

| Vital sign / visit | ICD<br>Mean (SE) | ILR<br>Mean (SE) | Mean difference<br>(95% CI) | P-value |
| --- | --- | --- | --- | --- |
| **Weight** |  |  |  |  |
| Month 6 | xx.xx (xx.xx) | xx.xx (xx.xx) | xx.xx (xx.xx, xx.x) |  |
| Month 12 | xx.xx (xx.xx) | xx.xx (xx.xx) | xx.xx (xx.xx, xx.x) |  |
| Month 18 | xx.xx (xx.xx) | xx.xx (xx.xx) | xx.xx (xx.xx, xx.x) |  |
| Etc. | xx.xx (xx.xx) | xx.xx (xx.xx) | xx.xx (xx.xx, xx.x) |  |
| Overall | xx.xx (xx.xx) | xx.xx (xx.xx) | xx.xx (xx.xx, xx.x) | 0.xxx |
| **BMI** |  |  |  |  |
| Month 6 | xx.xx (xx.xx) | xx.xx (xx.xx) | xx.xx (xx.xx, xx.x) |  |
| Month 12 | xx.xx (xx.xx) | xx.xx (xx.xx) | xx.xx (xx.xx, xx.x) |  |
| Month 18 | xx.xx (xx.xx) | xx.xx (xx.xx) | xx.xx (xx.xx, xx.x) |  |
| Etc. | xx.xx (xx.xx) | xx.xx (xx.xx) | xx.xx (xx.xx, xx.x) |  |
| Overall | xx.xx (xx.xx) | xx.xx (xx.xx) | xx.xx (xx.xx, xx.x) | 0.xxx |
| **SBP** |  |  |  |  |
| Month 6 | xx.xx (xx.xx) | xx.xx (xx.xx) | xx.xx (xx.xx, xx.x) |  |
| Month 12 | xx.xx (xx.xx) | xx.xx (xx.xx) | xx.xx (xx.xx, xx.x) |  |
| Month 18 | xx.xx (xx.xx) | xx.xx (xx.xx) | xx.xx (xx.xx, xx.x) |  |
| Etc. | xx.xx (xx.xx) | xx.xx (xx.xx) | xx.xx (xx.xx, xx.x) |  |
| Overall | xx.xx (xx.xx) | xx.xx (xx.xx) | xx.xx (xx.xx, xx.x) | 0.xxx |
| **DBP** |  |  |  |  |
| Month 6 | xx.xx (xx.xx) | xx.xx (xx.xx) | xx.xx (xx.xx, xx.x) |  |
| Month 12 | xx.xx (xx.xx) | xx.xx (xx.xx) | xx.xx (xx.xx, xx.x) |  |
| Month 18 | xx.xx (xx.xx) | xx.xx (xx.xx) | xx.xx (xx.xx, xx.x) |  |
| Etc. | xx.xx (xx.xx) | xx.xx (xx.xx) | xx.xx (xx.xx, xx.x) |  |
| Overall | xx.xx (xx.xx) | xx.xx (xx.xx) | xx.xx (xx.xx, xx.x) | 0.xxx |
| **Pulse rate** |  |  |  |  |
| Month 6 | xx.xx (xx.xx) | xx.xx (xx.xx) | xx.xx (xx.xx, xx.x) |  |
| Month 12 | xx.xx (xx.xx) | xx.xx (xx.xx) | xx.xx (xx.xx, xx.x) |  |
| Month 18 | xx.xx (xx.xx) | xx.xx (xx.xx) | xx.xx (xx.xx, xx.x) |  |
| Etc. | xx.xx (xx.xx) | xx.xx (xx.xx) | xx.xx (xx.xx, xx.x) |  |
| Overall | xx.xx (xx.xx) | xx.xx (xx.xx) | xx.xx (xx.xx, xx.x) | 0.xxx |
| **Respiratory rate** |  |  |  |  |
| Month 6 | xx.xx (xx.xx) | xx.xx (xx.xx) | xx.xx (xx.xx, xx.x) |  |
| Month 12 | xx.xx (xx.xx) | xx.xx (xx.xx) | xx.xx (xx.xx, xx.x) |  |
| Month 18 | xx.xx (xx.xx) | xx.xx (xx.xx) | xx.xx (xx.xx, xx.x) |  |
| Etc. | xx.xx (xx.xx) | xx.xx (xx.xx) | xx.xx (xx.xx, xx.x) |  |
| Overall | xx.xx (xx.xx) | xx.xx (xx.xx) | xx.xx (xx.xx, xx.x) | 0.xxx |

Notes: See Section 3.10 for details about the modelling approach. The overall model result will also be reported on the corresponding figure

**Table 9:** Analysis of quality of life scores.

| Score / visit | ICD<br>Mean (SE) | ILR<br>Mean (SE) | Mean difference<br>(95% CI) | P-value |
| --- | --- | --- | --- | --- |
| **MLHFQ Physical score** |  |  |  |  |
| Month 6 | xx.xx (xx.xx) | xx.xx (xx.xx) | xx.xx (xx.xx, xx.x) |  |
| Month 12 | xx.xx (xx.xx) | xx.xx (xx.xx) | xx.xx (xx.xx, xx.x) |  |
| Month 18 | xx.xx (xx.xx) | xx.xx (xx.xx) | xx.xx (xx.xx, xx.x) |  |
| Month 24 | xx.xx (xx.xx) | xx.xx (xx.xx) | xx.xx (xx.xx, xx.x) |  |
| Etc. | xx.xx (xx.xx) | xx.xx (xx.xx) | xx.xx (xx.xx, xx.x) |  |
| Overall | xx.xx (xx.xx) | xx.xx (xx.xx) | xx.xx (xx.xx, xx.x) | 0.xxx |
| **MLHFQ socio-economic score** |  |  |  |  |
| Month 6 | xx.xx (xx.xx) | xx.xx (xx.xx) | xx.xx (xx.xx, xx.x) |  |
| Month 12 | xx.xx (xx.xx) | xx.xx (xx.xx) | xx.xx (xx.xx, xx.x) |  |
| Month 18 | xx.xx (xx.xx) | xx.xx (xx.xx) | xx.xx (xx.xx, xx.x) |  |
| Month 24 | xx.xx (xx.xx) | xx.xx (xx.xx) | xx.xx (xx.xx, xx.x) |  |
| Etc. | xx.xx (xx.xx) | xx.xx (xx.xx) | xx.xx (xx.xx, xx.x) |  |
| Overall | xx.xx (xx.xx) | xx.xx (xx.xx) | xx.xx (xx.xx, xx.x) | 0.xxx |
| **MLHFQ emotional/psychological score** |  |  |  |  |
| Month 6 | xx.xx (xx.xx) | xx.xx (xx.xx) | xx.xx (xx.xx, xx.x) |  |
| Month 12 | xx.xx (xx.xx) | xx.xx (xx.xx) | xx.xx (xx.xx, xx.x) |  |
| Month 18 | xx.xx (xx.xx) | xx.xx (xx.xx) | xx.xx (xx.xx, xx.x) |  |
| Month 24 | xx.xx (xx.xx) | xx.xx (xx.xx) | xx.xx (xx.xx, xx.x) |  |
| Etc. | xx.xx (xx.xx) | xx.xx (xx.xx) | xx.xx (xx.xx, xx.x) |  |
| Overall | xx.xx (xx.xx) | xx.xx (xx.xx) | xx.xx (xx.xx, xx.x) | 0.xxx |
| **MLHFQ total score** |  |  |  |  |
| Month 6 | xx.xx (xx.xx) | xx.xx (xx.xx) | xx.xx (xx.xx, xx.x) |  |
| Month 12 | xx.xx (xx.xx) | xx.xx (xx.xx) | xx.xx (xx.xx, xx.x) |  |
| Month 18 | xx.xx (xx.xx) | xx.xx (xx.xx) | xx.xx (xx.xx, xx.x) |  |
| Month 24 | xx.xx (xx.xx) | xx.xx (xx.xx) | xx.xx (xx.xx, xx.x) |  |
| Etc. | xx.xx (xx.xx) | xx.xx (xx.xx) | xx.xx (xx.xx, xx.x) |  |
| Overall | xx.xx (xx.xx) | xx.xx (xx.xx) | xx.xx (xx.xx, xx.x) | 0.xxx |
| **EQ-5D VAS** |  |  |  |  |
| Month 6 | xx.xx (xx.xx) | xx.xx (xx.xx) | xx.xx (xx.xx, xx.x) |  |
| Month 12 | xx.xx (xx.xx) | xx.xx (xx.xx) | xx.xx (xx.xx, xx.x) |  |
| Month 18 | xx.xx (xx.xx) | xx.xx (xx.xx) | xx.xx (xx.xx, xx.x) |  |
| Month 24 | xx.xx (xx.xx) | xx.xx (xx.xx) | xx.xx (xx.xx, xx.x) |  |
| Etc. | xx.xx (xx.xx) | xx.xx (xx.xx) | xx.xx (xx.xx, xx.x) |  |
| Overall | xx.xx (xx.xx) | xx.xx (xx.xx) | xx.xx (xx.xx, xx.x) | 0.xxx |

Notes: See Sections 3.12.4 and 3.13.4 for details about the modelling approach. The overall model result will also be reported on the corresponding figure
MLHFQ = Minnesota Living with Heart Failure Questionnaire

**Table 10:** Summary of serious adverse events.

| Cause | ICD<br>(N= XXX) |  | ILR<br>(N= XXX) |  | Total<br>(N= XXX) |  | Fisher<br>exact<br>p-value |
| --- | --- | --- | --- | --- | --- | --- | --- |
|  | #events | N (%) | #events | N (%) | #events | N (%) |  |
| Serious adverse event | xxx | xxx (xx.x%) | xxx | xxx (xx.x%) | xxx | xxx (xx.x%) | 0.xxx |
| Death | xxx | xxx (xx.x%) | xxx | xxx (xx.x%) | xxx | xxx (xx.x%) |  |
| Life threatening illness or injury | xxx | xxx (xx.x%) | xxx | xxx (xx.x%) | xxx | xxx (xx.x%) |  |
| Persistent or permanent disability | xxx | xxx (xx.x%) | xxx | xxx (xx.x%) | xxx | xxx (xx.x%) |  |
| Inpatient hospitalisation | xxx | xxx (xx.x%) | xxx | xxx (xx.x%) | xxx | xxx (xx.x%) |  |
| Causing birth defect | xxx | xxx (xx.x%) | xxx | xxx (xx.x%) | xxx | xxx (xx.x%) |  |
| Requires medical or surgical intervention to prevent impairment to body structure or function | xxx | xxx (xx.x%) | xxx | xxx (xx.x%) | xxx | xxx (xx.x%) |  |

For each row, numbers represent the total number of events, the number and proportion of patients with at least one event of that type. The denominator is the number of patients randomised.

**Table 11:** Adverse events.

| Body system / event | ICD<br>(N= XXX) |  | ILR<br>(N= XXX) |  | Total<br>(N= XXX) |  | Fisher<br>exact<br>p-value |
| --- | --- | --- | --- | --- | --- | --- | --- |
|  | #events | N (%) | #events | N (%) | #events | N (%) |  |
| **Body system 1** | xxx | xxx (xx.x%) | xxx | xxx (xx.x%) | xxx | xxx (xx.x%) | **0.xxx** |
| Event 1 | xxx | xxx (xx.x%) | xxx | xxx (xx.x%) | xxx | xxx (xx.x%) |  |
| Event 2 | xxx | xxx (xx.x%) | xxx | xxx (xx.x%) | xxx | xxx (xx.x%) |  |
| Etc. | xxx | xxx (xx.x%) | xxx | xxx (xx.x%) | xxx | xxx (xx.x%) |  |
| **Body system 2** | xxx | xxx (xx.x%) | xxx | xxx (xx.x%) | xxx | xxx (xx.x%) | **0.xxx** |
| Event 1 | xxx | xxx (xx.x%) | xxx | xxx (xx.x%) | xxx | xxx (xx.x%) |  |
| Event 2 | xxx | xxx (xx.x%) | xxx | xxx (xx.x%) | xxx | xxx (xx.x%) |  |
| Etc. | xxx | xxx (xx.x%) | xxx | xxx (xx.x%) | xxx | xxx (xx.x%) |  |
| **Body system 3** | xxx | xxx (xx.x%) | xxx | xxx (xx.x%) | xxx | xxx (xx.x%) | **0.xxx** |
| Event 1 | xxx | xxx (xx.x%) | xxx | xxx (xx.x%) | xxx | xxx (xx.x%) |  |
| Event 2 | xxx | xxx (xx.x%) | xxx | xxx (xx.x%) | xxx | xxx (xx.x%) |  |
| Etc. | xxx | xxx (xx.x%) | xxx | xxx (xx.x%) | xxx | xxx (xx.x%) |  |
| Etc. |  |  |  |  |  |  |  |

*Note: Order body systems, and events within each body system, by decreasing order of total frequency*
*For each row, numbers represent the total number of events, the number and proportion of patients with at least one event of that type. The denominator is the number of patients randomised.*

**Table 12:** Cause of death.

| **Cause** | **ICD**<br>**(N= XXX)**<br>**N (%)** | **ILR**<br>**(N= XXX)**<br>**N (%)** | **Total**<br>**(N= XXX)**<br>**N (%)** |
| --- | --- | --- | --- |
| Cause 1 | XXX (XX.X%) | XXX (XX.X%) | XXX (XX.X%) |
| Cause 2 | XXX (XX.X%) | XXX (XX.X%) | XXX (XX.X%) |
| Cause 3 | XXX (XX.X%) | XXX (XX.X%) | XXX (XX.X%) |
| Etc... | XXX (XX.X%) | XXX (XX.X%) | XXX (XX.X%) |

*Note: order by decreasing order of total frequency. The denominator is the number of patients randomised.*

**Table 13:** Hospitalisations.

| **Cause** | **ICD**<br>**(N= XXX)**<br>**#events N (%)** | **ILR**<br>**(N= XXX)**<br>**#events N (%)** | **Total**<br>**(N= XXX)**<br>**#events N (%)** |
| --- | --- | --- | --- |
| Cause 1 | XXX XXX<br>(XX.X%) | XXX XXX<br>(XX.X%) | XXX XXX<br>(XX.X%) |
| Cause 2 | XXX XXX<br>(XX.X%) | XXX XXX<br>(XX.X%) | XXX XXX<br>(XX.X%) |
| Cause 3 | XXX XXX<br>(XX.X%) | XXX XXX<br>(XX.X%) | XXX XXX<br>(XX.X%) |
| Etc... | XXX XXX<br>(XX.X%) | XXX XXX<br>(XX.X%) | XXX XXX<br>(XX.X%) |

*Note: order by decreasing order of total frequency*
*For each hospitalisation, numbers represent the total number of events, the number and proportion of patients with at least one hospitalisation of that type. The denominator is the number of patients randomised.*

- Demographic characteristics: age (years), sex, height (cm), weight (kg), BMI (kg/m2)
- Race
- Ethnicity
- Smoking status
- Cause of LV dysfunction (ischaemic LV dysfunction or non-ischaemic LV dysfunction i.e. DCM)
- Cardiovascular history and risk factors (e.g. diabetes, hypertension, etc.)
- Systolic blood pressure, diastolic blood pressure
- Serum creatinine (µmol/L)
- Medical history including baseline medications

### 3.8 Concomitant medications

Concomitant medications will be reported as the number and proportion of patients taking a medication at least once over the course of the study as well as separately at each visit. This will be done by category of medication (e.g. beta-blockers, renin–angiotensin system inhibitors, etc.).

### 3.9 Compliance and protocol deviations

Intervention details will be summarised as the number and proportion of patients in each arm who receive the allocated intervention as well as those who cross-over i.e. patients randomised to ILR who receive an ICD and patients randomised to the ICD arm who either do not receive or had the ICD deactivated or removed.

Protocol deviations will be summarised as the number of deviations by type and classification. All protocol deviations will be listed together with a description of the deviation. No formal statistical test will be applied.

### 3.10 Analysis of vital signs

The following vital signs are collected at baseline, month 3, month 6, month 12 and every 12 months thereafter: Weight, BMI, SBP, DBP, pulse rate and respiratory rate.

Vital signs will be summarised graphically by plotting means and 95% confidence intervals by visit and treatment arm. Differences between arms will be estimated using repeated-measure linear mixed models with a normal distribution and identity link. The model will include all post-randomization values after reallocation of the EOS visit (see Section 3.5). Fixed effects will include treatment, visit, treatment by visit interaction term as well as the value of the outcome at baseline included as a continuous variable. Within-patient correlations will be modelled using a repeated patient effect assuming a compound-symmetry (exchangeable) structure. A random site effect will be included to account for stratification by site. The overall effect of the intervention will be estimated as the mean difference and 95% CI between the intervention and control arm over the entire follow-up i.e. by combining data obtained across all visits. The effect of the intervention at specific time points (e.g. at Month 12) will be estimated from the same model using contrasts; however, no formal statistical test will be applied to individual visits.

### 3.11 Analysis of the primary outcome

#### 3.11.1 Main analysis

The primary endpoint, time from randomisation to first occurrence of sudden cardiac death or haemodynamically significant ventricular arrhythmia, will be summarised using cumulative incidence functions and analysed using a Fine and Gray model, [5] where deaths other than sudden cardiac deaths will be considered as competing risks. The randomised arm will be included as a fixed variable with no other covariates included. The effect of the intervention will be estimated as a hazard ratio (HR) and its 95% confidence interval (CI) obtained from the Fine and Gray model of the sub-distribution hazard. The analysis will be censored at the time when a patient was last known to be alive and free of event. As a sensitivity analysis, a Cox model will determine the cause-specific HR, which estimates the risk of SCD or haemodynamically significant ventricular arrhythmia in subjects who are still alive.

#### 3.11.2 Adjusted analyses

Adjusted analyses will be performed by adding the following covariates to the main model: age (continuous), sex (male vs female), heart failure aetiology (ICM or DCM) and baseline left ventricular ejection fraction (30-40 vs >40-50). The adjusted treatment effect will be reported as the HR and 95% CI.

#### 3.11.3 Subgroup analyses

Six pre-specified subgroup analyses will be carried out irrespective of whether there is a significant treatment effect on the primary outcome. Subgroups are defined as follows:

- Age (<70 vs 70 or more)
- Sex (Male vs Female)
- LVEF (36-<40, 40-<46, 46-50)
- Type of cardiomyopathy (ICM vs DCM)
- NYHA class at baseline (I vs II-III)
- QRS duration (130 ms and < 130 ms)

The analysis for each subgroup will be performed by adding the subgroup variable as well as its interaction with the intervention as fixed effects to the main analysis model (see Section 3.11.1). Within each subgroup, summary measures will include raw counts and percentages within each treatment arm, as well as the hazard ratio for treatment effect with a 95% confidence interval. The results will be displayed on a forest plot including the p-value for heterogeneity corresponding to the interaction term between the intervention and the subgroup variable.

#### 3.11.4 Treatment of missing data

Given the nature of the primary endpoint, there is no reasonable way to assess whether some events are potentially missing (i.e. unreported). We will therefore perform the analysis assuming that all events that occurred were reported and without imputation of missing data. All reported events will be included in the time-to-first-event analysis with every participant censored at the time when the participant was last known to be alive and free of an event and with non-sudden deaths treated as a competing risk. See Section 3.11.1 for more details about the main analysis. The same approach will be used for every secondary survival endpoint.

### 3.12 Analyses of secondary outcomes

#### 3.12.1 Sudden cardiac death

Time to sudden cardiac death will be analysed using the same approach as the primary outcome described in Section 3.11.1. The main analysis will consist of a survival analysis of time to first event treating deaths that are not sudden cardiac deaths as a competing risk using a Fine and Gray model. A supporting analysis will be a Cox “cause-specific” survival model estimating the risk of sudden cardiac death in patients who are still alive. No additional adjusted analyses or subgroup analyses will be applied.

#### 3.12.2 Haemodynamically significant ventricular arrhythmia

Time to HSVA will be analysed using the same approach as the primary outcome described in Section 3.11.1. The main analysis will consist of a survival analysis of time to first event treating all deaths as a competing risk using a Fine and Gray model. A supporting analysis will be a Cox “cause-specific” survival model estimating the risk of HVA in patients who are still alive. No additional adjusted analyses or subgroup analyses will be applied.

Details of HVA events including the type (i.e. monomorphic ventricular tachycardia, polymorphic ventricular tachycardia or ventricular fibrillation) and duration (more or less than 30 seconds) will be reported descriptively as the number of events and the number and proportion of patients experiencing each event category at least once.

#### 3.12.3 All-cause mortality

All-cause mortality will be analysed as time between randomisation and death using a survival analysis censored at the time when the patient was last known to be alive. A Kaplan-Meier plot will be used to describe survival rates. Differences in survival will be tested using a Cox model with the randomised arm as a fixed covariate.

#### 3.12.4 Quality of life assessed by Minnesota Living with Heart Failure Questionnaire

For each of the 3 domains, we will calculate a score. The score of each domain as well as the overall score will be summarised graphically by plotting means and 95% confidence intervals by visit and treatment arm. For the total score, differences between arms will be estimated using repeated-measure linear mixed models with a normal distribution and identity link. The model will include all post-randomization values after reallocation of the EOS visit (see Section 3.5). Fixed effects will include treatment, visit, treatment by visit interaction term as well as the value of the outcome at baseline included as a continuous variable. Within-patient correlations will be modelled using a repeated patient effect assuming a compound-symmetry (exchangeable) structure. A random site effect will be included to account for stratification by site. The overall effect of the intervention will be estimated as the mean difference and 95% CI between the intervention and control arm over the entire follow-up i.e. by combining data obtained across all visits. The effect of the intervention at specific time points (e.g. at Month 12) will be estimated from the same model using contrasts; however, no formal statistical test will be applied to individual visits. No modelling or formal tests will be applied to the sub-domain scores.

#### 3.12.5 Heart-failure related hospitalisations

Time to heart-failure related hospitalisation including heart-failure deaths will be analysed using the same approach as the primary outcome described in Section 3.11.1. The main analysis will consist of a survival analysis of time to first event treating deaths that are not related to heart failure as a competing risk using a Fine and Gray model. A supporting analysis will be a Cox “cause-specific” survival model estimating the risk of heart-failure related hospitalisation in patients who are still alive. No additional adjusted analyses or subgroup analyses will be applied

#### 3.12.6 Cardiovascular mortality

Time to cardiovascular death will be analysed using the same approach as the primary outcome described in Section 3.11.1. The main analysis will consist of a survival analysis of time to first event treating deaths that are not due to a cardiovascular cause as a competing risk using a Fine and Gray model. A supporting analysis will be a Cox “cause-specific” survival model estimating the risk of cardiovascular death in patients who are still alive. No additional adjusted analyses or subgroup analyses will be applied..

### 3.13 Analysis of safety and other/exploratory outcomes

#### 3.13.1 Sustained ventricular arrhythmia

Time to sustained ventricular arrhythmia will be analysed using the same approach as the primary outcome described in Section 3.11.1. The main analysis will consist of a survival analysis of time to first event treating all deaths as a competing risk using a Fine and Gray model. A supporting analysis will be a Cox “cause-specific” survival model estimating the risk of sustained VA in patients who are still alive. No additional adjusted analyses or subgroup analyses will be applied.

#### 3.13.2 Cardiovascular hospitalisations

Time to cardiovascular hospitalisation including cardiovascular deaths will be analysed using the same approach as the primary outcome described in Section 3.11.1. The main analysis will consist of a survival analysis of time to first event treating deaths that are not due to a cardiovascular cause as a competing risk using a Fine and Gray model. A supporting analysis will be a Cox “cause-specific” survival model estimating the risk of cardiovascular hospitalisation or death in patients who are still alive. No additional adjusted analyses or subgroup analyses will be applied.

#### 3.13.3 Change in New York Heart Association (NYHA) functional class

NYHA class is collected at baseline, month 3, month 6, month 12 and every 12 months thereafter. It consists of 4 ordinal categories from 1 to 4. The number and proportion of participants in each class at each visit will be described using tables and stacked bar charts. Changes in NYHA class will be analysed as time to worsening defined as moving to a worse class than the baseline class (i.e. from class I to class II or more, from class II to class III or more, or from class III to class IV). It will be analysed using a survival analysis of time to first event (increase in class) treating death as a competing risk using a Fine and Gray model. A supporting analysis will be a Cox “cause-specific” survival model estimating the risk of NYHA increase in patients who are still alive. No additional adjusted analyses or subgroup analyses will be applied to this outcome.

#### 3.13.4 Quality of life assessed by the EQ-5D-5L Questionnaire

Each of the five dimensions will be summarised descriptively over time (i.e. by visit) and treatment arm using stacked bar charts. The VAS score will be summarised graphically using box plots by visit and treatment arm. Differences between arms will be estimated using repeated-measure linear mixed models with a normal distribution and identity link. The model will include all post-randomization values after reallocation of the EOS visit (see Section 3.5). Fixed effects will include treatment, visit, treatment by visit interaction term as well as the value of the outcome at baseline included as a continuous variable. Within-patient correlations will be modelled using a repeated patient effect assuming a compound-symmetry (exchangeable) structure. A random site effect will be included to account for stratification by site. The overall effect of the intervention will be estimated as the mean difference and 95% CI between the intervention and control arm over the entire follow-up i.e. by combining data obtained across all visits. The effect of the intervention at specific time points (e.g. at Month 12) will be estimated from the same model using contrasts; however, no formal statistical test will be applied to individual visits.

For Australian sites, a detailed economic evaluation using the EQ-5D index will be conducted after publication of the main results. Details will be outlined in a separate document.

#### 3.13.5 Adverse events

Adverse events will be summarised by severity and relationship to the intervention. This will be done overall and by category of event (body system and preferred term). For each category, we will report the total number of events as well as the number of patients with at least one event. Proportions of patients with adverse events will be compared between treatment arms using Fisher’s exact test, both overall and by body system. No test will be done at the preferred term level.

#### 3.13.6 Hospitalisations

Hospitalisations will be reported descriptively by type as the total number of events as well as the number and percentage of patients experiencing each type at least once. No formal test will be applied.

#### 3.13.7 Causes of deaths

Deaths will be reported descriptively by cause as the number and percentage of patients. No formal test will be applied.

### 3.14 Win ratio analysis

Components of the primary outcome and key secondary outcomes will be analysed jointly using a win ratio approach which ranks all outcomes in order of importance [6]. The following hierarchy will be used:

1. Time to sudden cardiac death
2. Time to haemodynamically significant ventricular arrhythmia
3. Number of heart failure hospitalisations
4. Total Minnesota Living with Heart Failure Questionnaire score at 24 months

Every participant randomised to the ICD arm will be paired with every participant randomised to the ILR arm. For illustrative purposes, if we had 130 patients randomised to one arm and 127 to the other, we would compare a total of 16,510 pairs (130 × 127). Within each pair, we will compare outcomes in a hierarchical fashion until a winner has been determined. If no winner can be declared after comparing all outcomes, the pair will be tied. We will start by comparing all-cause mortality to determine a “winner”. The winner will be the participant with the longest survival time. If neither participant died and a winner cannot be declared on mortality alone, we will then proceed to the next step and compare time to haemodynamically significant ventricular arrhythmia. As with death, the winner will be the participant with the longest time to first event. If neither patient experienced the event of interest or both experienced the event on the same day, the pair will be declared a tie and we will proceed to the next outcome in the hierarchy. Only events that occur during the time that both participants are in the study will be used (e.g. if one participant withdraws after 1 year, only the events occurring in the first year will be used to compare the pair). The approach will proceed in a stepwise fashion, moving to the next outcome in the hierarchy, until a decision has been reached for every pair. The full decision process is described in the table below. The win ratio will then be calculated as the proportion of “ICD winners” (the participant in the ICD arm had a better outcome) divided by the proportion of “ILR winners” (the participant in the ILR arm had a better outcome)

To complete the interpretation, we will also compute the win odds and net benefit [7]. Calculations will be performed using the R package WINS developed by Cui and Huang [8].

| Step | Outcome | Decision rule |
| --- | --- | --- |
| 1 | Time to death | The winner is the patient with the longest survival time. If neither died or if both died on the same day, go to step 2. |
| 2 | Time to first haemodynamically significant ventricular arrhythmia | The winner is the patient with the longest time to event. If neither patient experienced the event or both patients had it on the same day, go to step 3. |
| 3 | Number of heart failure hospitalisations | The winner is the patient with the lowest number of events. If neither patient experienced the event or both patients experienced the same number of events, go to step 4. |
| 4 | Total Minnesota Living with Heart Failure Questionnaire score at 24 months | The winner is the patient with the lowest (best) score at the 24-month visit. If both patients have the same score or if one or more patients has missing data, the pair is considered a tie. |

An “incremental analysis” will also be performed by calculating the win ratio and 95% CI after adding each outcome i.e. for the first outcome only, for the first two together, the first three, and so on (cf Figure 10 for details).

**Figure 1:** Consort flowchart. Consort flow chart showing the number of subjects at each step (e.g. screened, randomised, lost to follow up and included in the primary outcome analysis)

**Figure 2.** <vital sign> by visit. Programming notes:

- Do this figure for all vital signs listed in Section 3.10
- Show means and 95%CI. Display raw means on the graph as numbers near each dot and denominators below the x-axis.
- Also display overall mean difference, 95% confidence interval and p-value from repeated-measure linear mixed model (see Section 3.10 for details)

**Figure 3:** Cumulative incidence curve of time to <outcome>. Programming notes:

- Do this figure for all primary and secondary outcomes of a survival nature
- Add number at risk every 12 months
- Display median and quartiles as well as results from the survival model used for the main analysis (hazard ratio, 95% CI and p-value). See Section 3.11.1 for details of the main analysis model.
- Except for death, the results should come from a Fine and Gray model where death from a cause that is not part of the outcome definition is treated as a competing risk (See Section 3.11.1 for details of the main analysis model)

**Figure 4:**
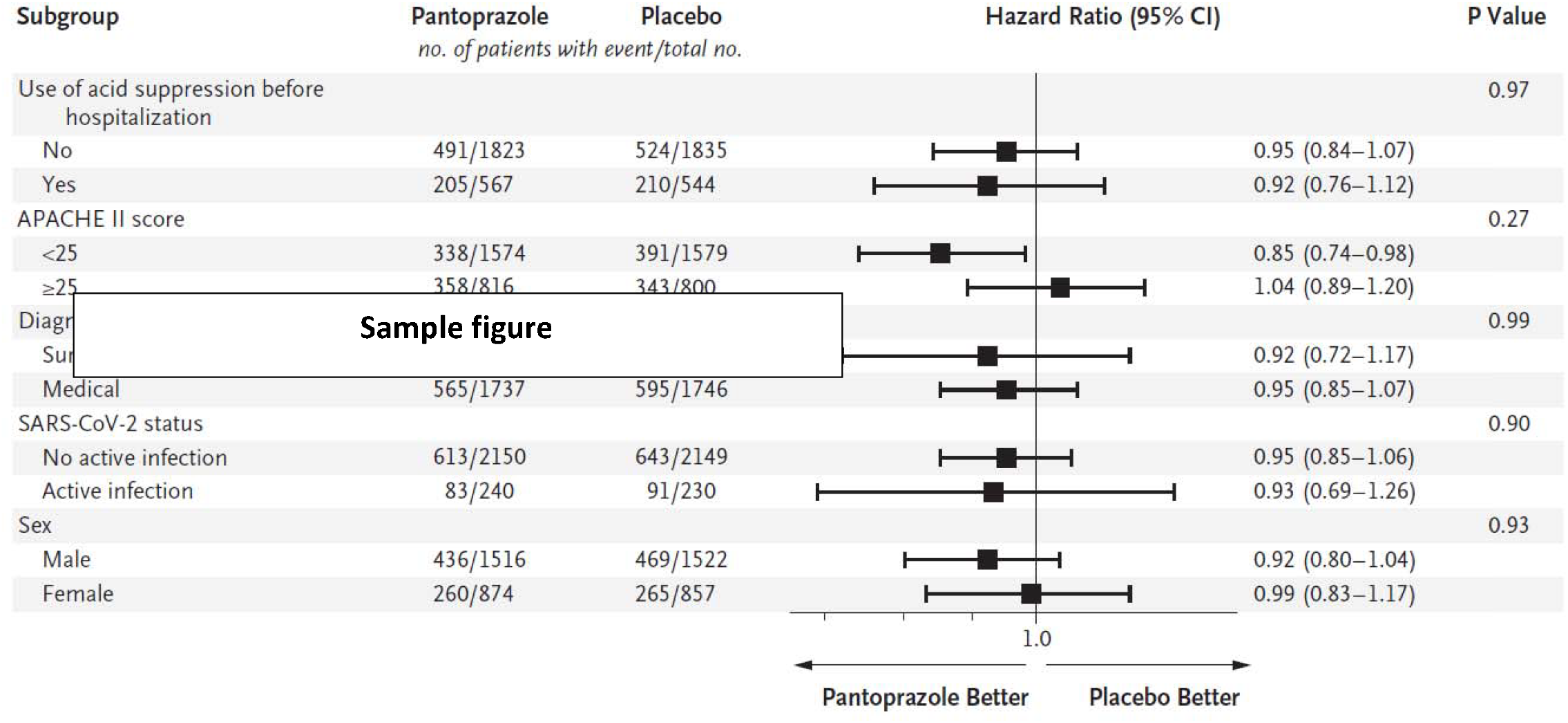
Forest plot for subgroup analysis of primary outcome. Forest plot showing the effect of ICD vs ILR on the primary outcome (time to first occurrence of SCD or haemodynamically significant ventricular arrhythmia) by pre-specified baseline subgroups. Display p-value for heterogeneity corresponding to the interaction term between the intervention and the subgroup variable. Here is an example: Notes: Do for the primary outcome only. Figure above for illustrative purposes only (source: https://www.nejm.org/doi/abs/10.1056/NEJMoa2404245)

**Figure 5.** Stacked bar chart of New York Heart Association (NYHA) class by visit. Programming notes:

- NYHA class is collected at baseline, month 3, month 6, month 12 and every 12 months thereafter. It consists of 4 ordinal categories from 1 to 4.
- Create a stacked bar chart with two columns (one for each treatment group) for each visit including baseline and, within each column, stack the categories from 1 to 5 and colour code using a gradient (e.g. white to dark red).
- Display raw percentages within categories and denominators below the x-axis

**Figure 6.** Stacked bar chart of <EQ-5D question> by visit. Programming notes: • Do this figure for each of the 5 EQ-5D questions • Create a stacked bar chart with two columns (one for each treatment group) for each visit including baseline and, within each column, stack the categories from 1 to 5 and colour code using a gradient (e.g. white to dark red). • Display raw percentages within categories and denominators below the x-axis

**Figure 7:** Minnesota Living with Heart Failure <domain/total> score by visit. Programming notes: • Do this figure for each of the 3 domains as well as the overall score (see Section 3.12.4) • Show means and 95%CI. Display raw means on the graph as numbers near each dot and denominators below the x-axis. • Also display overall mean difference, 95% confidence interval and p-value from repeated-measure linear mixed model (see Section 3.10 for details)

**Figure 8:** Boxplot of EQ-5D VAS score by visit. Programming notes: • Create a boxplot for the VAS score by visit and treatment arm • Display denominators below the x-axis • Display overall mean difference, 95% confidence interval and p-value from repeated-measure linear mixed model (see Section 3.13.4 for details)

**Figure 9.**
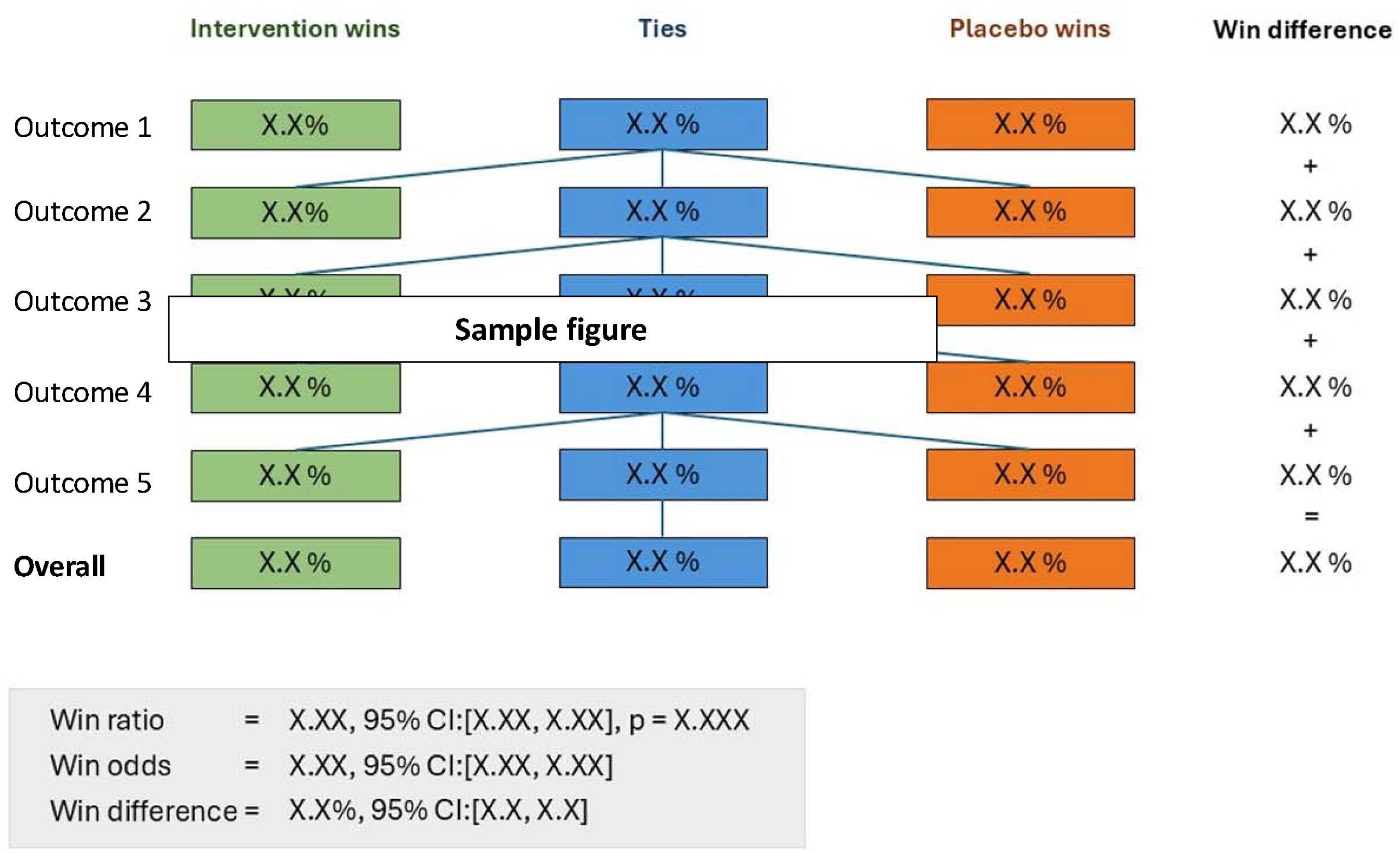
Win ratio decision tree.

**Figure 10.**
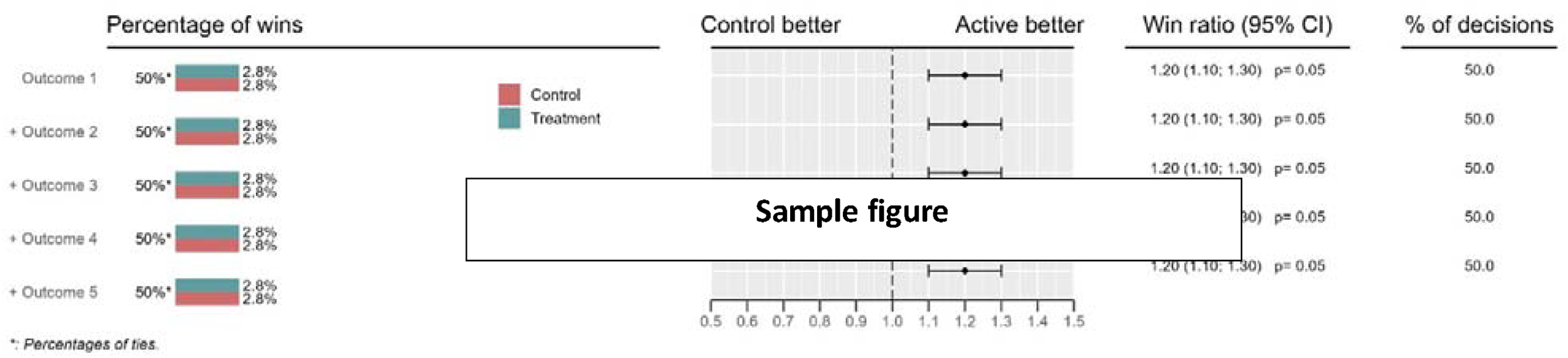
Win ratio cumulative analysis.

## Data Availability

No relevant

## APPENDIX 1 Proposed Tables and figures

## Notes

### Competing Interest Statement

The authors have declared no competing interest.

### Clinical Trial

NCT01918215

### Funding Statement

The study is funded by:
- Biotronik Australia Pty Ltd
- Bayer Australia Ltd

### Author Declarations

The following ethical approvals were obtained: Southern Adelaide Clinical Human Research Ethics Committee - Host institution: Joint partnership, Southern Adelaide Local Health Network and Flinders University - Approval number: 430.14 (HREC/14/SAC/449) University of Tasmania Human Research Ethics Committee - Host institution: University of Tasmania - Approval number: H0016170 Sir Charles Gairdner and Osborne Park Health Care Group Human Research Ethics Committee - Host institution: WA Department of Health - Approval number: RGS0000001765 WA Health Central HREC (new central HREC for WA sites effective 7 May 2025) - Host institution: WA Department of Health - Approval number: RGS0000001765 (same as above migrated from SCGOPHCG to WA Central HREC) Ethics Committee State Chamber of Physicians of Baden Wurttemberg - Host institution: State Chamber of Physicians of Baden Wurttemberg - Approval number: F-2015-095-RS South East Scotland Research Ethics Committee 01 - Host institution: National Health Service Lothian (Lothian NHS Board) - Approval number: 16/SS/0153

### Summary of Updates

Corrected mistake in definition of sustained VA

